# Risk Factors for Suicidal Behavior in Youth and the Impact of SARS-CoV-2 Infection: A Retrospective Case-Control Study

**DOI:** 10.1101/2024.12.02.24318197

**Authors:** Kathleen P. Heslin, Michelle Montero, Stephen V. Faraone, Yanli Zhang-James

## Abstract

**Background:** Suicide and self-harm remain critical concerns in youth. This study compares patients with and without suicidality or self-harm (SOSH), suicidality (SI/SA), and COVID-19 to investigate 53 pre-existing risk factors associated with suicidality in patients with and without COVID-19.

**Methods:** A retrospective case-control study was conducted using TriNetX data from 111,631,250 patients across 78 healthcare networks. This study included patients aged 0-21 with any healthcare visit between January 20, 2020, and May 11, 2023.

**Outcomes:** Comparison groups shared many risk factors, with specific differences. Children with SOSH and COVID-19 had higher odds of support group problems, personality disorder, thyroid disorders, and insomnia; children with SOSH without COVID-19 had higher odds of upbringing problems, anxiety and nonpsychotic disorders, sleep disorders, and autism. Adolescents with SOSH and COVID-19 had higher odds of parent-child conflict; adolescents with SOSH without COVID-19 had higher odds of education and literacy problems. Children with SI/SA and COVID-19 had higher odds of support group problems, personality disorders, and asthma; children with SI/SA without COVID-19 had higher odds of autism. Adolescents with SI/SA and COVID-19 had higher odds of asthma. The effect size of COVID-19 was not significant. SOSH was associated with increased odds of prior SARS-CoV-2 infection in children (OR 2.42) and adolescents (OR 1.88).

**Interpretation:** This study confirms known SOSH risk factors and demonstrates their association with suicidality. We observed a significant association between SOSH and preceding SARS-CoV-2 infection. This underscores the need to focus on suicide risk in youth affected by COVID-19.

## INTRODUCTION

Over the past decade, suicidal ideation and suicide attempt have increased in children and adolescents^(1)^. Suicide remains a leading cause of death in youth^(2)^, and presentations to medical facilities for emergency care related to suicide have increased nearly five-fold over the past decade^(3)^. Since the start of the COVID-19 pandemic, there has been interest in the effect of the pandemic and SARS-COV-2 infection on suicidality. Hospital presentations for suicide attempt (SA), self-harm (SH), and suicidal ideation (SI) increased during the pandemic^(4–6)^. A smaller number of studies suggest the opposite, including decreased admissions for suicidal behavior during the pandemic lockdown^(7)^. Despite much research about suicidality or self-harm (SOSH) related to the pandemic, little research has assessed the relationship between SOSH and SARS-COV-2 infection. In adults, an association has been observed between SARS-COV-2 and suicidal ideation or suicidality both before^(8)^ and after^(9)^ SARS-COV-2 infection; and between self-harm or thoughts of self-harm and history of SARS-COV-2 infection^(10)^. Our recent study of the electronic health record (EHR) data of 13 million youth found that COVID-19 is a significant risk factor for suicidal ideation and attempt, with hazard ratios 2·2 (2·0-2·4) and 2·8 (2·0-3·8), respectively, in adolescents even two years after their initial SARS-CoV-2 infection^(11)^.

Suggested etiologies include the potential association of suicidality and inflammation^(12)^. Although COVID-19 is no longer a public health crisis, the virus remains prevalent in the United States, and associated long-term psychiatric outcomes remain pertinent, particularly in youth. Large studies using real-world data to evaluate the impact of SARS-COV-2 infection itself on suicidal behavior in children and adolescents are urgently needed.

In addition to COVID-19, many risk factors for suicidality have been investigated extensively during the past 50 years^(13)^. These include psychiatric disorders like ADHD and conduct disorder^(5)^ and symptoms like anxiety ^(14, 15)^; socioeconomic factors including low-income or sexual minority status^(14)^; and medical diagnoses like asthma^(16, 17)^ and diabetes^(18, 19)^. A meta-analysis showed that across 50 years, much of the research has been homogeneously focused on risk factors from the five main categories^(13)^, few studies examined the combination of multiple risk factors, and few used large real-world data. Furthermore, the predictive accuracies from these studies have been mediocre^(13)^. Establishing valid and predictive risk factors is the first step toward improving the prevention and treatment of suicide. Such insights will guide future research in developing more accurate predictive models, identifying novel predictors, and most importantly avoiding wasted effort on non-informative risk factors.

To achieve this goal, we used a large de-identified global database of electronic health records from 13 million children and adolescents to comprehensively assess all the previously identified risk factors for suicidality, specifically those which we could map to the electronic health record data, in relation to SARS-CoV-2 using a retrospective case-control design.

## METHODS

We conducted a retrospective case-control study using TriNetX, a global research network of de-identified data from electronic medical records (EMRs). This study focused on the Research Network, a collection of data from 111,631,250 patients across 78 healthcare networks (HCOs). This study population included 13,038,353 patients 0-21 who had any healthcare visit within the study period of January 20, 2020 (the first reported COVID-19 case in the United States [US]) until May 11, 2023 (the end of the US federal COVID-19 Public Health Emergency). The study used de-identified patient data and was exempted from institutional review.

### STUDY DESIGN

The study population included children (0-12) and adolescents (13-21). In each age group, we compared characteristics (pre-existing psychiatric, medical, and socioeconomic factors) between populations of patients diagnosed with first-time COVID-19 who later developed suicidality or self-harm (SOSH), and patients diagnosed with first-time COVID-19 who did not later develop SOSH.

### COVID-19 CASE CRITERIA

We defined COVID-19 positive cases based on either positive laboratory tests for SARS-CoV-2 or COVID-19 diagnostic codes (see Supplement) within the study period. Exclusion criteria included patients diagnosed with other viral diseases (ICD B25-B24) or other specified viral diseases (B33·8) in the study period, and patients with a personal history of COVID-19 (Z86·16). In this text, “COVID-19” refers to both COVID-19 diagnosis and acute SARS-CoV-2 infection.

### SUICIDALITY AND SELF-HARM (SOSH)

*SOSH* was defined as the presence of any of the following ICD-10 codes: suicide attempt (suicide attempt, initial encounter [T14·91XA]; or suicide attempt [T14·91]); suicidal ideations (R45·851); or intentional self-harm (X71-X83).

### SUICIDALITY (SI/SA)

Suicidality (SI/SA) was defined as the presence of any of the following: suicide attempt (suicide attempt, initial encounter [T14·91XA]; or suicide attempt [T14·91]); or suicidal ideations (R45·851).

### COHORT DESIGN

For the primary analysis, in each age group (0-12, 13-21), we created four cohorts: patients who, within the study period, had been diagnosed with COVID-19 and were diagnosed with SOSH after their COVID-19 diagnosis (COVID+, SOSH+); patients with a COVID-19 diagnosis who were not later diagnosed with SOSH (COVID+,SOSH-); patients who developed SOSH but did not have COVID-19 during the study period (COVID-, SOSH+); and patients who were not diagnosed with COVID-19 or SOSH in the study period (COVID-, SOSH-).

### POPULATION COMPARISON

In the “Compare Outcomes” function, we selected 53 psychiatric, medical, and socioeconomic characteristics (“pre-existing characteristics”) associated with suicidality or self-harm in previous literature (see Table 1). For all group comparisons, we examined the presence of these characteristics before the index event. Index event was defined as any healthcare visit that occurred during the study period.

**Table 1.**
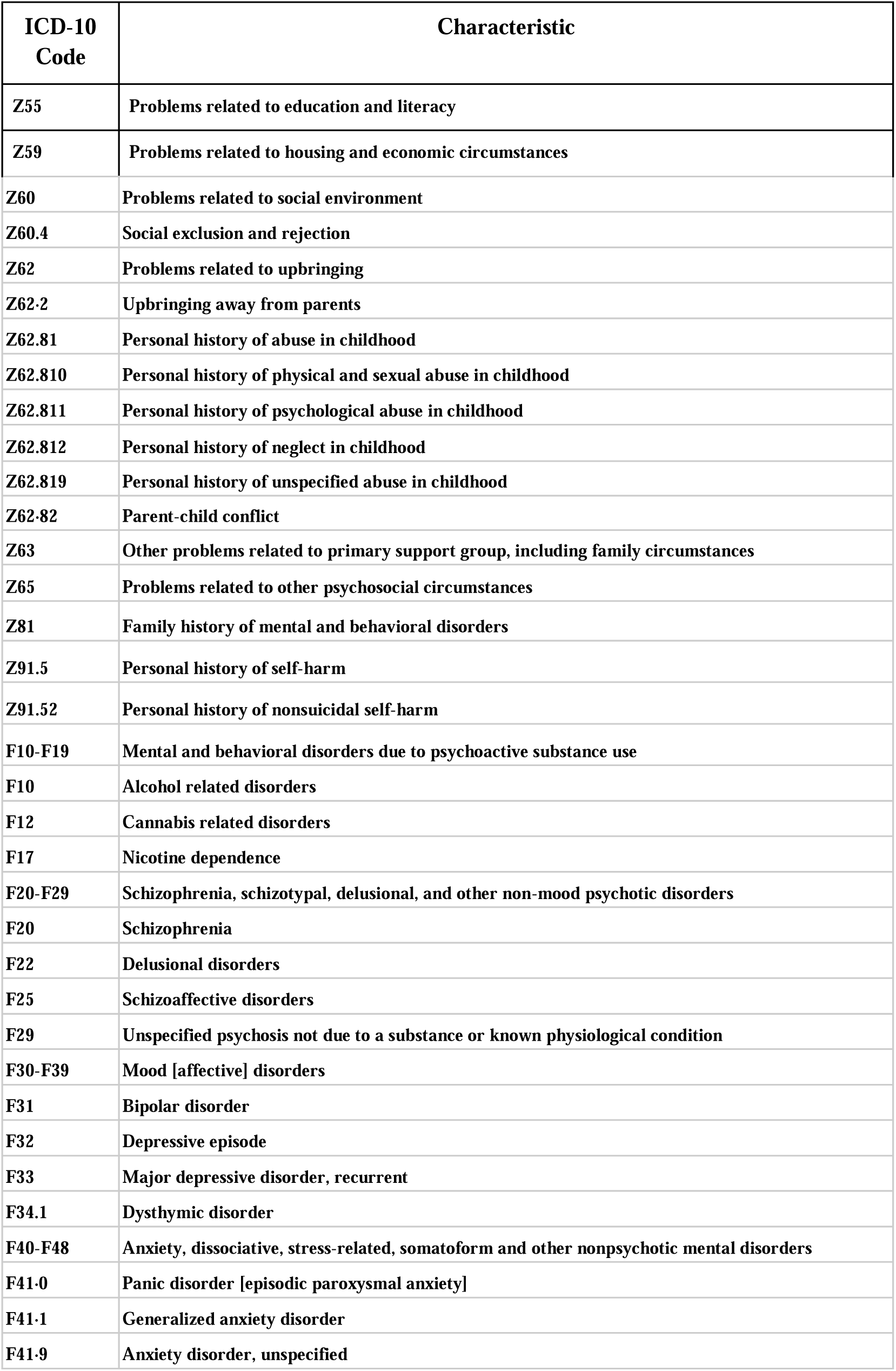
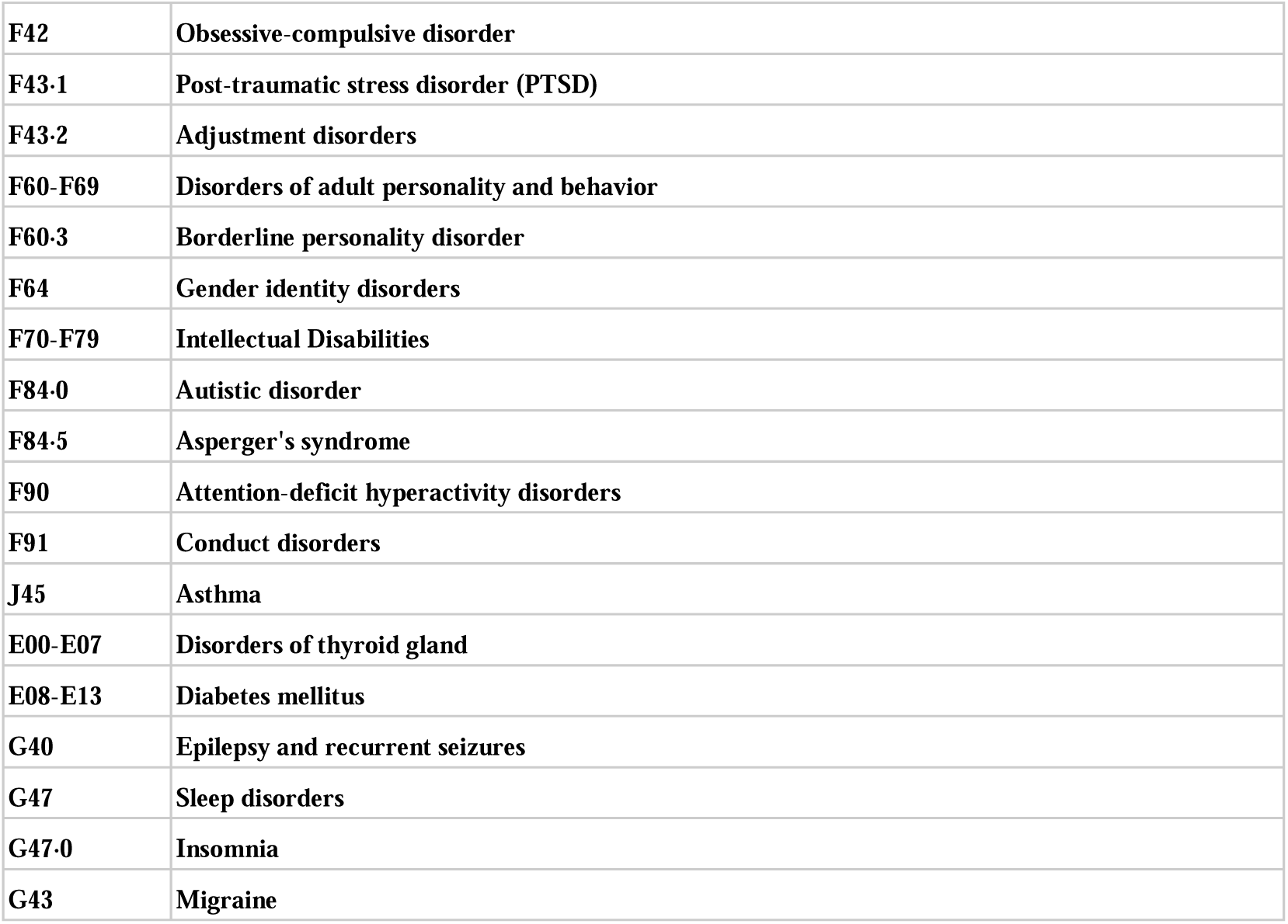
Socioeconomic, psychiatric, and medical characteristics assessed for all comparison groups.

| ICD-10 Code | Characteristic |
| --- | --- |
| Z55 | Problems related to education and literacy |
| Z59 | Problems related to housing and economic circumstances |
| Z60 | Problems related to social environment |
| Z60.4 | Social exclusion and rejection |
| Z62 | Problems related to upbringing |
| Z62.2 | Upbringing away from parents |
| Z62.81 | Personal history of abuse in childhood |
| Z62.810 | Personal history of physical and sexual abuse in childhood |
| Z62.811 | Personal history of psychological abuse in childhood |
| Z62.812 | Personal history of neglect in childhood |
| Z62.819 | Personal history of unspecified abuse in childhood |
| Z62.82 | Parent-child conflict |
| Z63 | Other problems related to primary support group, including family circumstances |
| Z65 | Problems related to other psychosocial circumstances |
| Z81 | Family history of mental and behavioral disorders |
| Z91.5 | Personal history of self-harm |
| Z91.52 | Personal history of nonsuicidal self-harm |
| F10-F19 | Mental and behavioral disorders due to psychoactive substance use |
| F10 | Alcohol related disorders |
| F12 | Cannabis related disorders |
| F17 | Nicotine dependence |
| F20-F29 | Schizophrenia, schizotypal, delusional, and other non-mood psychotic disorders |
| F20 | Schizophrenia |
| F22 | Delusional disorders |
| F25 | Schizoaffective disorders |
| F29 | Unspecified psychosis not due to a substance or known physiological condition |
| F30-F39 | Mood [affective] disorders |
| F31 | Bipolar disorder |
| F32 | Depressive episode |
| F33 | Major depressive disorder, recurrent |
| F34.1 | Dysthymic disorder |
| F40-F48 | Anxiety, dissociative, stress-related, somatoform and other nonpsychotic mental disorders |
| F41-0 | Panic disorder [episodic paroxysmal anxiety] |
| F41-1 | Generalized anxiety disorder |
| F41-9 | Anxiety disorder, unspecified |
| **F42** | **Obsessive-compulsive disorder** |
| **F43-1** | **Post-traumatic stress disorder (PTSD)** |
| **F43-2** | **Adjustment disorders** |
| **F60-F69** | **Disorders of adult personality and behavior** |
| **F60-3** | **Borderline personality disorder** |
| **F64** | **Gender identity disorders** |
| **F70-F79** | **Intellectual Disabilities** |
| **F84-0** | **Autistic disorder** |
| **F84-5** | **Asperger's syndrome** |
| **F90** | **Attention-deficit hyperactivity disorders** |
| **F91** | **Conduct disorders** |
| **J45** | **Asthma** |
| **E00-E07** | **Disorders of thyroid gland** |
| **E08-E13** | **Diabetes mellitus** |
| **G40** | **Epilepsy and recurrent seizures** |
| **G47** | **Sleep disorders** |
| **G47-0** | **Insomnia** |
| **G43** | **Migraine** |

### PROPENSITY SCORE-MATCHING

Using TriNetX’s propensity score matching (PSM) algorithm, we balanced cohorts based on ten demographic factors: age at index event, sex (male/female), race (American Indian or Alaska Native, Asian, Black or African American, Native Hawaiian or Other Pacific Islander, and white), and ethnicity (Hispanic or Latino, Not Hispanic or Latino). The PSM algorithm uses “greedy nearest neighbor matching” with a caliper set at 0·1 pooled standard deviations for individual patients in the smaller cohort to find best matches from the larger cohort. Combined matched cohorts were incorporated into our comparison analysis following the validation of propensity scores.

### ANALYSIS

For our primary analysis, using the TriNetX “Compare Outcomes” function, we compared the four cohorts four ways: COVID-19(+), SOSH(+) vs. COVID-19(+), SOSH(-); COVID-19(+), SOSH(+) vs. COVID-19(-), SOSH(+); COVID-19(+), SOSH(+) vs. COVID-19(-), SOSH(-); and COVID-19(-), SOSH(+) vs. COVID-19(-), SOSH(-). We compared these cohorts after propensity score-matching based on pre-existing demographic factors at the time of index event. During each comparison, if fewer than 10 patients in either cohort were associated with a characteristic, then that characteristic was excluded from analysis. In TriNetX, populations smaller than 10 are listed as “10” to protect patient privacy and are not reliable for analysis^(20)^.

Where the primary analysis compared cohorts based on development of SOSH, we repeated the analysis on smaller cohorts to focus on suicidality alone. The second analysis focused on suicidal ideations and suicide attempt (SI/SA). We carried out the following comparisons: COVID-19 (+), SI/SA(+) vs. COVID-19 (+), SI/SA(-); COVID-19 (+), SI/SA(+) vs. COVID-19(-), SI/SA(+); COVID-19 (+), SI/SA(+) vs. COVID-19 (-), SI/SA(-); COVID-19 (-), SI/SA(+) vs. COVID-19 (-), SI/SA(-). We repeated the procedures from the SOSH analysis (population comparison, propensity score-matching, and cohort comparison).

### STATISTICAL ANALYSIS

For our primary and secondary analyses, we computed odds ratios and 95% confidence intervals (CI), Z-scores, and p values according to Altman and Bland^(21)^. We used the Bonferroni method of correction with an adjusted alpha level of p<0·0009 based on a total of 53 tests. We determined the degree of COVID-19 effect by calculating the variance off the odds ratios, based on Altman and Bland^(21)^ and Higgins and Thomas^(22)^, as described in the supplementary information.

## RESULTS

In the TriNetX Research Network, a total of 7,341,636 children and 5,696,717 adolescents had a visit in the study period. Among these, 454,758 children and 374,567 adolescents had COVID-19 infection. In the total population, 9,901 children (0·13%) and 124,387 adolescents (2·18%) reported suicide ideation, intentional self-harm, or suicide attempt (SOSH). Among patients with COVID-19,1375 children (0·30%) and 14845 adolescents (3·96%) had SOSH. In the general population, 9540 children (0·13%) and 120001 adolescents (2·11%) had SI/SA. Among patients with COVID-19, 1311 children (0·29%) and 15521 adolescents (4·14%) had SI/SA.

After matching, in the 0-12 group, patients were 50·84% male, 49·15% female, with an average age of 8. This cohort was 57·33% white, 25·72% Black, 1·032% Asian, 0·74% American Indian or Alaska Native, 0·74% Native Hawaiian or Pacific Islander, 14·15% Hispanic, and 71·26% non-Hispanic. In the 13-21 group, patients were 32·03% male, 67·87% female, with an average age of 14. Patients were 60·63% white, 22·29% Black, 2·13% Asian, 0·47% American Indian or Alaska Native, 0·35% Native Hawaiian or Pacific Islander, 11·96% Hispanic, and 70·7% non-Hispanic.

Compared to patients without SOSH, patients with SOSH had greater odds of a preceding diagnosis of COVID-19, for both children (OR 2·42, 95%CI (2·29, 2·56), p<0·0008) and adolescents (OR 1·88, 95%CI (1·85, 1·92), p<0·0008). Compared to patients without SI/SA, patients with SI/SA also had greater odds of a preceding diagnosis of COVID-19, both for children (OR 2·32, 95%CI (2·19, 2·46), p<0·0008) and adolescents (OR 1·90, 95% CI (1·86, 1·93), p<0·0008).

### Association with Suicidality and Self-Harm in COVID-19(-) Patients

#### SOSH and SI/SA

##### Ages 0-12

Among patients with no diagnosis of COVID-19 before or during the study period, patients with SOSH had a significantly greater odds of pre-existing psychiatric diagnosis (ADHD (OR 6·71, 95%CI (4·87, 9·24), p<0·0008), conduct disorders (OR 7·68, 95%CI (4·83, 12·21), p<0·0008), anxiety and nonpsychotic disorders (OR 6·82, 95%CI (4·94, 9·43), p<0·0008), unspecified anxiety (OR 8·43, 95%CI (5·06, 14·05), p<0·0008), autistic disorder (OR 3·83, 95%CI (2·30, 6·40), p<0·0008)), medical diagnoses (sleep disorders (OR 2·01, 95%CI (1·45, 2·80), p<0·0008)), and one socioeconomic diagnosis (problems related to upbringing (OR 8·08, 95%CI (4·29, 15·22), p<0·0008)) (Table 2) (Figure 1).

**Figure 1.**
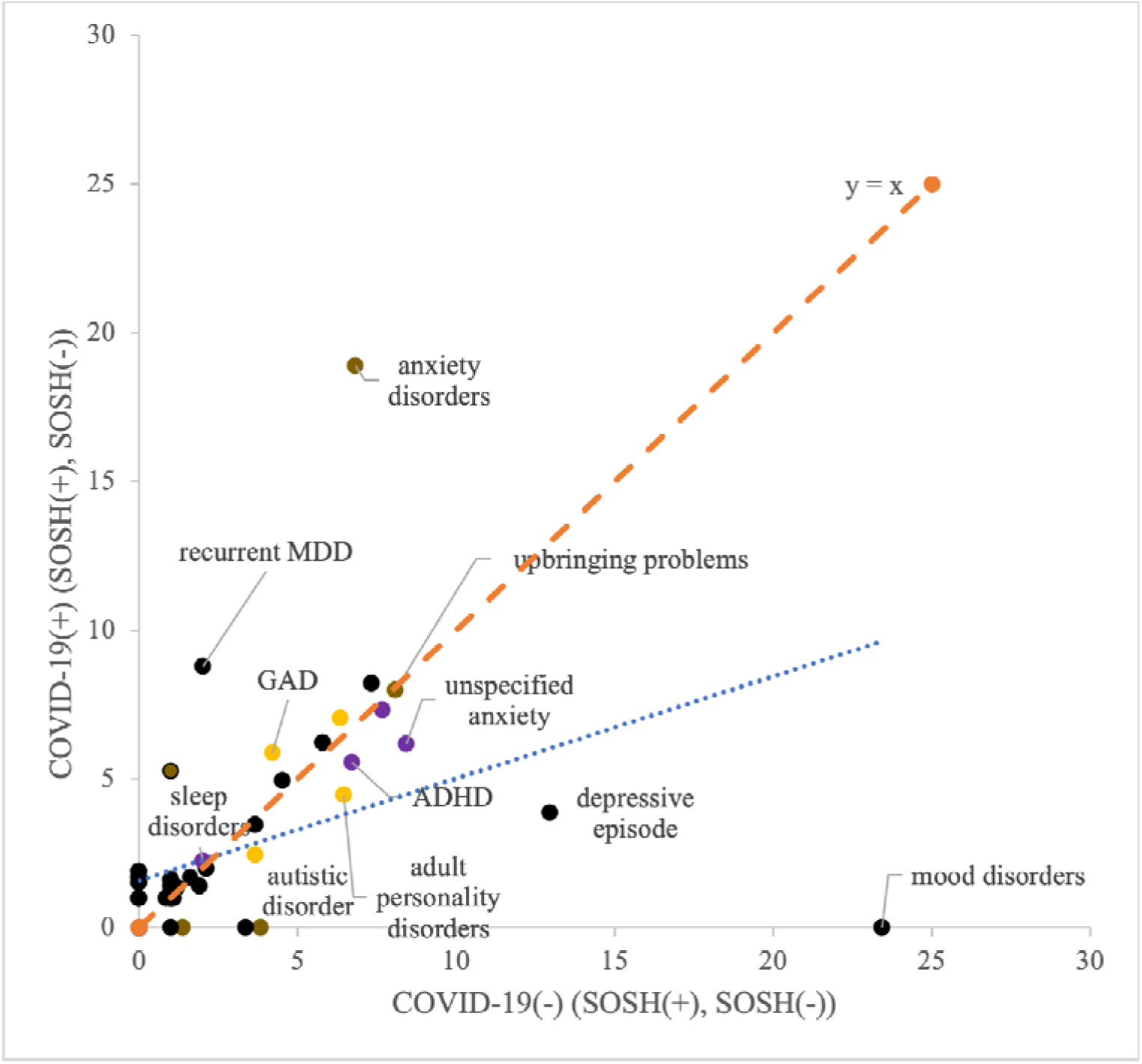
Comparison of odds ratios of medical/psychiatric/socioeconomic characteristics in patients 0-12 years old with or without suicide attempt/suicidal ideation/self-injurious behavior in COVID-19 (-) and COVID-19 (+) patients. Labeled characteristics are a sample of the significant findings. Key: Purple denotes significance in both COVID-19(+) and COVID-19(-) patients; yellow denotes significance in only COVID-19(+) patients; brown denotes significance in only COVID-19(-) patients; and black denotes a lack of significance in each group.

**Table 2.** Prevalence and odds ratios of medical/psychiatric/socioeconomic characteristics in patients 0-12 with or without SOSH among COVID-19 (-) patients. Statistical comparisons are represented by odds ratios (OR) with 95% confidence intervals (CI). Significant p-values are indicated with an asterisk (*).

| **ICD-10 Code** | **Characteristic** | **COVID-19(-), SOSH(+)** |  | **COVID-19(-), SOSH(-)** |  | **OR** | **95%CI** | **P value** |
| --- | --- | --- | --- | --- | --- | --- | --- | --- |
|  |  | **N** | **%** | **N** | **%** |  |  |  |
| **SOCIOECONOMIC DIAGNOSES** |  |  |  |  |  |  |  |  |
| **Z62** | **Problems related to upbringing** | 84 | 6.20% | 11 | 0.81% | 8.08 | (4.29, 15.22) | *$<0.0008$ |
| **PSYCHIATRIC DIAGNOSES** |  |  |  |  |  |  |  |  |
| **F40-F48** | **Anxiety, dissociative, stress-related, somatoform and other nonpsychotic mental disorders** | 262 | 19.31% | 46 | 3.39% | 6.82 | (4.94, 9.43) | *$<0.0008$ |
| **F41-9** | **Anxiety disorder, unspecified** | 131 | 9.65% | 17 | 1.25% | 8.43 | (5.06, 14.05) | *$<0.0008$ |
| **F84-0** | **Autistic disorder** | 70 | 5.16% | 19 | 1.40% | 3.83 | (2.30, 6.40) | *$<0.0008$ |
| **F90** | **Attention-deficit hyperactivity disorders** | 263 | 19.38% | 47 | 3.46% | 6.71 | (4.87, 9.24) | *$<0.0008$ |
| **F91** | **Conduct disorders** | 146 | 10.76% | 21 | 1.55% | 7.68 | (4.83, 12.21) | *$<0.0008$ |
| **MEDICAL DIAGNOSES** |  |  |  |  |  |  |  |  |
| J45 | Asthma | 151 | 11.13% | 113 | 8.32% | 1.38 | (1.07, 1.78) | 0.0138 |
| G40 | Epilepsy and recurrent seizures | 19 | 1.40% | 22 | 1.62% | 0.86 | (0.46, 1.60) | 0.6517 |
| G47 | Sleep disorders | 110 | 8.11% | 57 | 4.20% | 2.01 | (1.45, 2.80) | *<0.0008 |
| G43 | Migraine | 14 | 1.03% | 14 | 1.03% | 1.00 | (0.48, 2.11) | 0.9986 |

Among this COVID-19 negative population, patients with SI/SA had a similar profile: higher odds of psychiatric diagnoses (ADHD (OR 9·04, 95%CI (6·27, 13·06), p<0·0008), conduct disorders (OR 10·82, 95%CI (6·53, 17·95), p<0·0008), adjustment disorders (OR 6·77, 95%CI (3·57, 12·85), p<0·0008), anxiety disorders (OR 8·61, 95%CI (6·02, 12·32), p<0·0008), unspecified anxiety (OR 9·44, 95%CI (5·50, 16·21), p<0·0008), autistic disorder (OR 4·30, 95%CI (2·47, 7·46), p<0·0008)) and one medical diagnosis (sleep disorders (OR 2·00, 95%CI (1·41, 2·83), p=0·0001)) (Table 3) (Figure 2).

**Figure 2.**
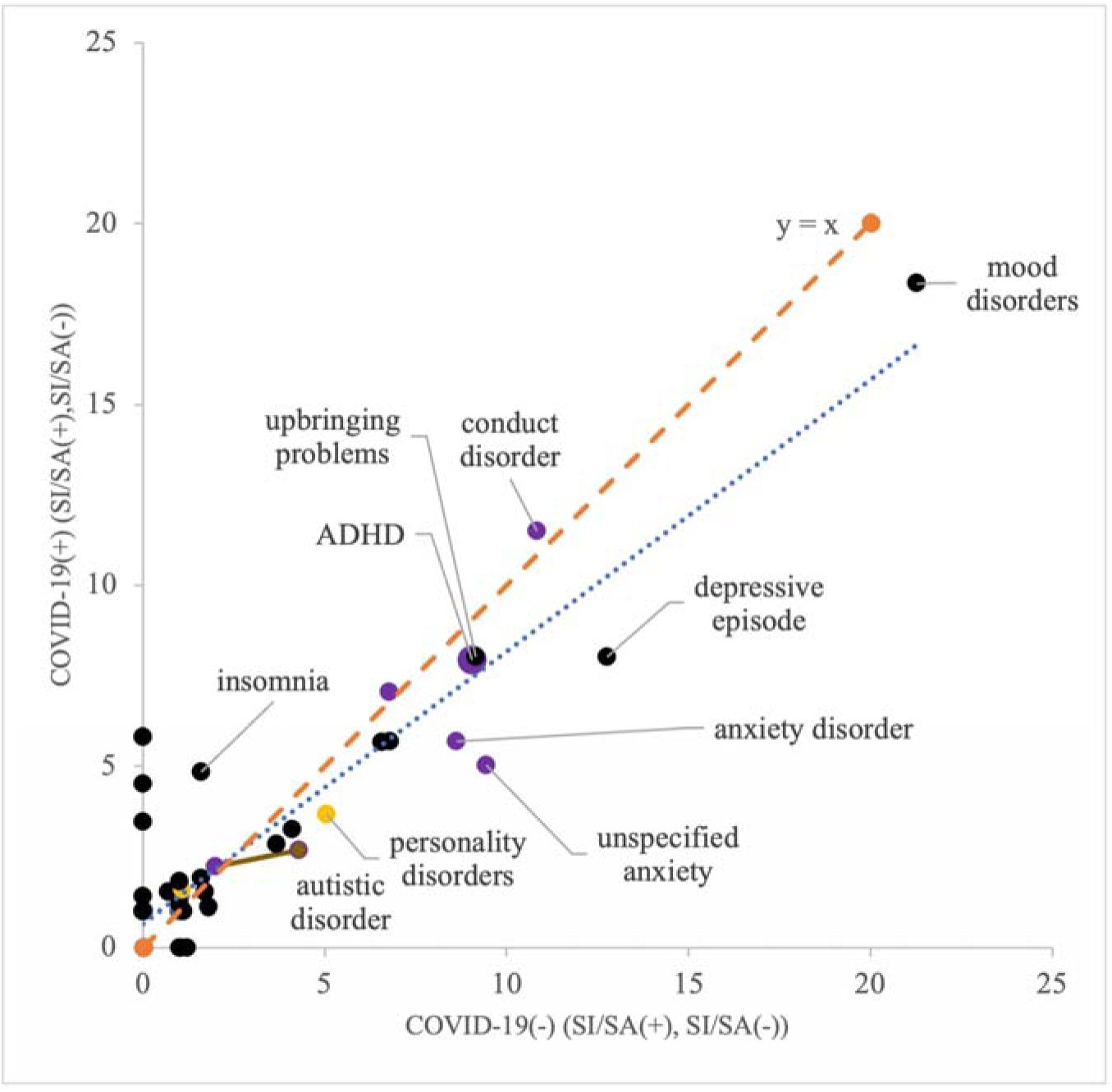
Comparison of odds ratios of medical/psychiatric/socioeconomic characteristics in patients 0-12 years old with or without suicide attempt/suicidal ideation in COVID-19 (-) and COVID-19 (+) patients. Labeled characteristics are a sample of the significant findings. Key: Purple denotes significance in both COVID-19(+) and COVID-19(-) patients; yellow denotes significance in only COVID-19(+) patients; brown denotes significance in only COVID-19(-) patients; and black denotes a lack of significance in each group.

**Table 3.** Prevalence and odds ratios of medical/psychiatric/socioeconomic characteristics in patients 0-12 with or without SI/SA among COVID-19 (-) patients. Statistical comparisons are represented by odds ratios (OR) with 95% confidence intervals (CI). Significant p-values are indicated with an asterisk (*).

| ICD-10 Code | Characteristic | COVID-19(-), SI/SA(+) |  | COVID-19(-), SI/SA(-) |  | OR | 95%CI | P value |
| --- | --- | --- | --- | --- | --- | --- | --- | --- |
|  |  | N | % | N | % |  |  |  |
| **PSYCHIATRIC DIAGNOSES** |  |  |  |  |  |  |  |  |
| F40-F48 | Anxiety, dissociative, stress-related, somatoform and other nonpsychotic mental disorders | 256 | 19.65% | 36 | 2.76% | 8.61 | (6.02, 12.32) | *<0.0008 |
| F41.9 | Anxiety disorder, unspecified | 129 | 9.90% | 15 | 1.15% | 9.44 | (5.50, 16.21) | *<0.0008 |
| F43.2 | Adjustment disorders | 71 | 5.45% | 11 | 0.84% | 6.77 | (3.57, 12.85) | *<0.0008 |
| F84.0 | Autistic disorder | 66 | 5.07% | 16 | 1.23% | 4.30 | (2.47, 7.46) | *<0.0008 |
| F90 | Attention-deficit hyperactivity disorders | 254 | 19.49% | 34 | 2.61% | 9.04 | (6.27, 13.06) | *<0.0008 |
| F91 | Conduct disorders | 163 | 12.51% | 17 | 1.30% | 10.82 | (6.53, 17.95) | *<0.0008 |
| **MEDICAL DIAGNOSES** |  |  |  |  |  |  |  |  |
| J45 | Asthma | 122 | 9.36% | 115 | 8.82% | 1.07 | (0.82, 1.40) | 0.6419 |
| G40 | Epilepsy and recurrent seizures | 12 | 0.92% | 17 | 1.30% | 0.70 | (0.33, 1.48) | 0.3598 |
| G47 | Sleep disorders | 98 | 7.52% | 51 | 3.91% | 2.00 | (1.41, 2.83) | *0.0001 |

##### Ages 13-21

Adolescents without COVID-19 who developed SOSH had a significantly greater odds of pre-existing psychiatric risk factors like ADHD (OR 2·02, 95%CI (1·80, 2·27), p<0·0008), conduct disorders (OR 2·60, 95%CI (2·19,3·08), p<0·0008), adjustment disorders (OR 1·90, 95%CI (1·50,2·42), p<0·0008), PTSD (OR 6·27, 95%CI (3·76,10·47), p<0·0008), bipolar disorder (OR 4·01, 95%CI (2·23,7·21), p<0·0008), disorders of adults personality and behavior (OR 2·49, 95%CI (2·00,3.11), p<0·0008), borderline personality disorder (OR 4·09, 95%CI (2·23,7·50), p<0·0008), mood disorders (OR 4·07, 95%CI ( 3·37,4·92), p<0·0008), anxiety and nonpsychotic disorders (OR 2·29, 95%CI (2·02,2·60), p<0·0008), unspecified anxiety disorder (OR 2·44, 95%CI (2·05,2·89), p<0·0008), generalized anxiety disorder (OR 2·27, 95%CI (1·75,2·96), p<0·0008), depressive episode (OR 3·75, 95%CI (2·98,4·71), p<0·0008), recurrent major depressive disorder (OR 7·62, 95%CI (4·17,13·92), p<0·0008), and autistic disorder (OR 1·63, 95%CI (1·26, 2·11), p=0·0002) (Table 4) (Figure 3). These patients also had higher odds of socioeconomic factors: problems related to primary support group (OR 2·15, 95%CI (1·53,3·01), p<0·0008), problems related to education and literacy (OR 2·20, 95%CI (1·52,3·19), p<0·008), problems related to upbringing (OR 3·04, 95%CI (2·15,4·30), p<0·0008), family history of mental and behavioral disorders (OR 4·71, 95%CI (2·97,7·46), p<0·0008), and upbringing away from parents (OR 3·13, 95%CI (1·78,5·50), p<0·0008). There were increased odds of only two medical diagnoses: sleep disorders (OR 1·37, 95%CI (1·18,1·59), p<0·0008) and insomnia (OR 2.03, 95%CI (1·44,2·86), p<0·0008).

**Figure 3.**
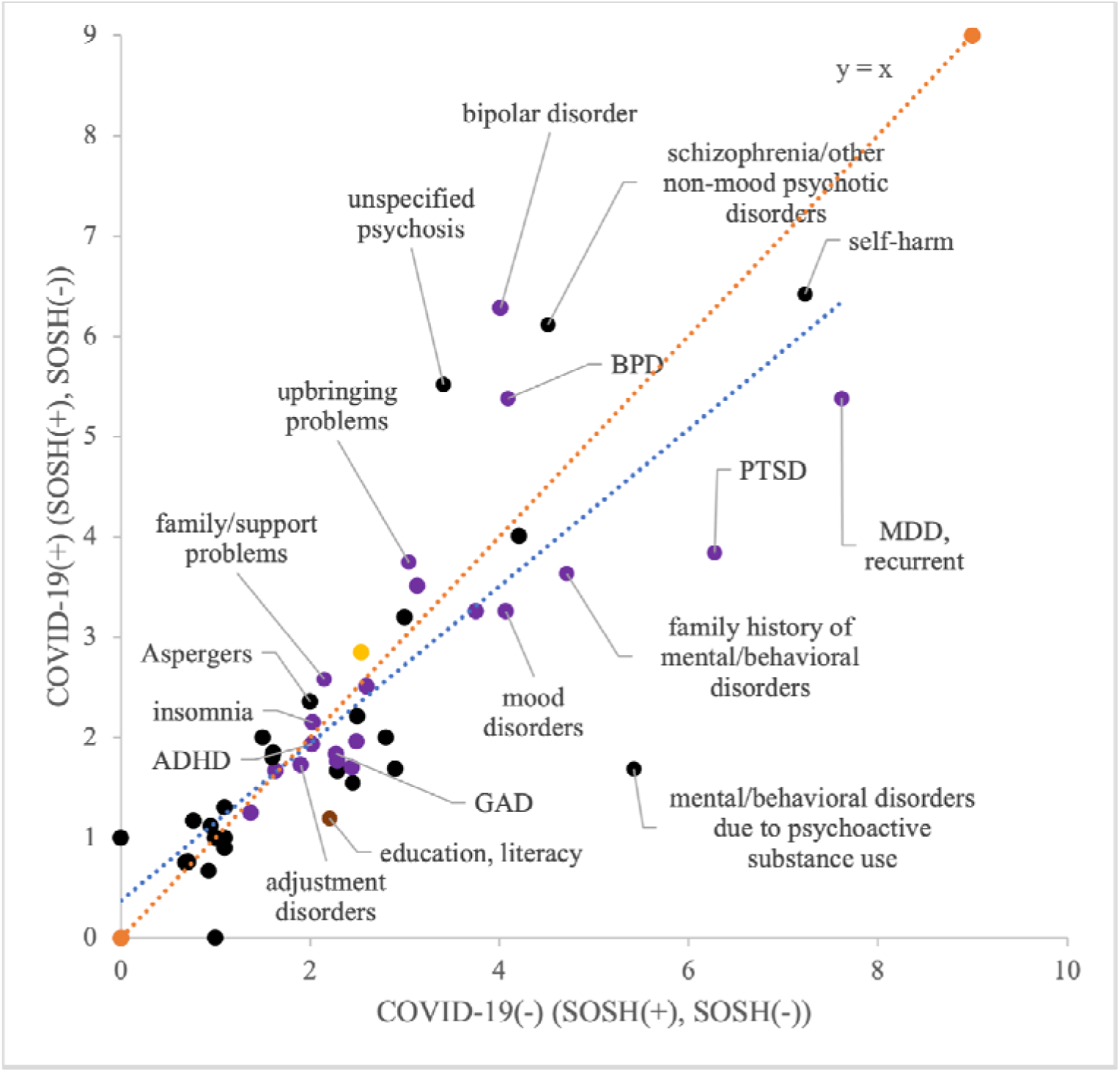
Comparison of odds ratios of medical/psychiatric/socioeconomic characteristics in patients 13-21 years old with or without suicide attempt/suicidal ideation/self-injurious behavior in COVID-19 (-) and COVID-19 (+) patients. Labeled characteristics are a sample of the significant findings. Key: Purple denotes significance in both COVID-19(+) and COVID-19(-) patients; yellow denotes significance in only COVID-19(+) patients; brown denotes significance in only COVID-19(-) patients; and black denotes a lack of significance in each group.

**Table 4.**
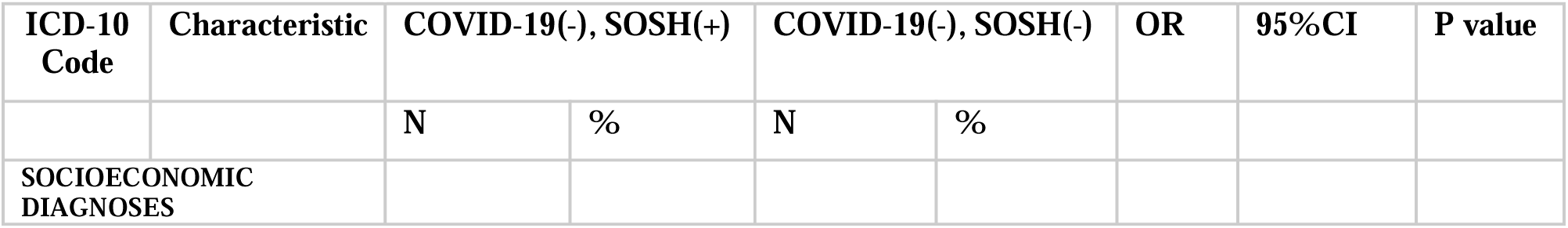
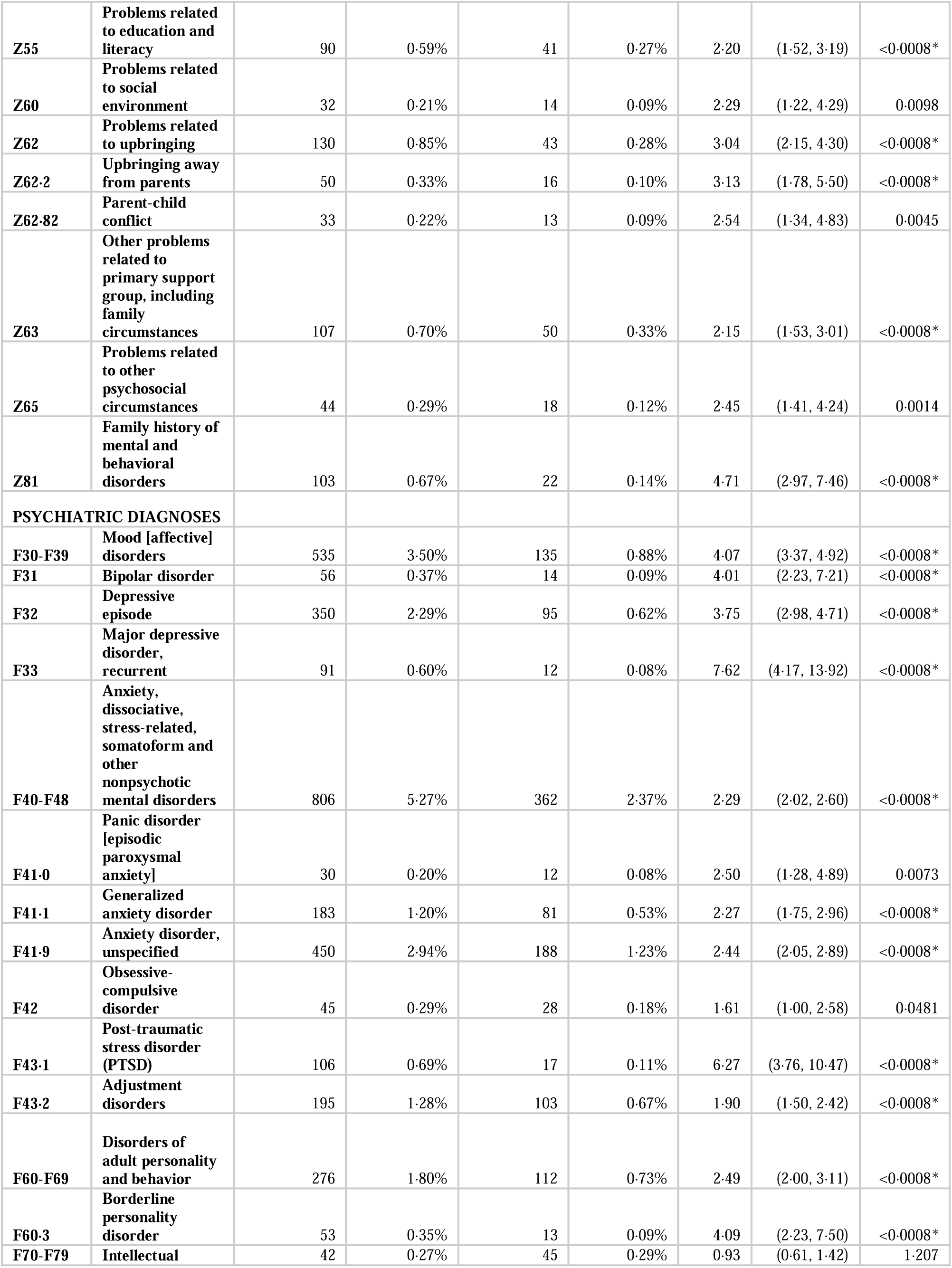
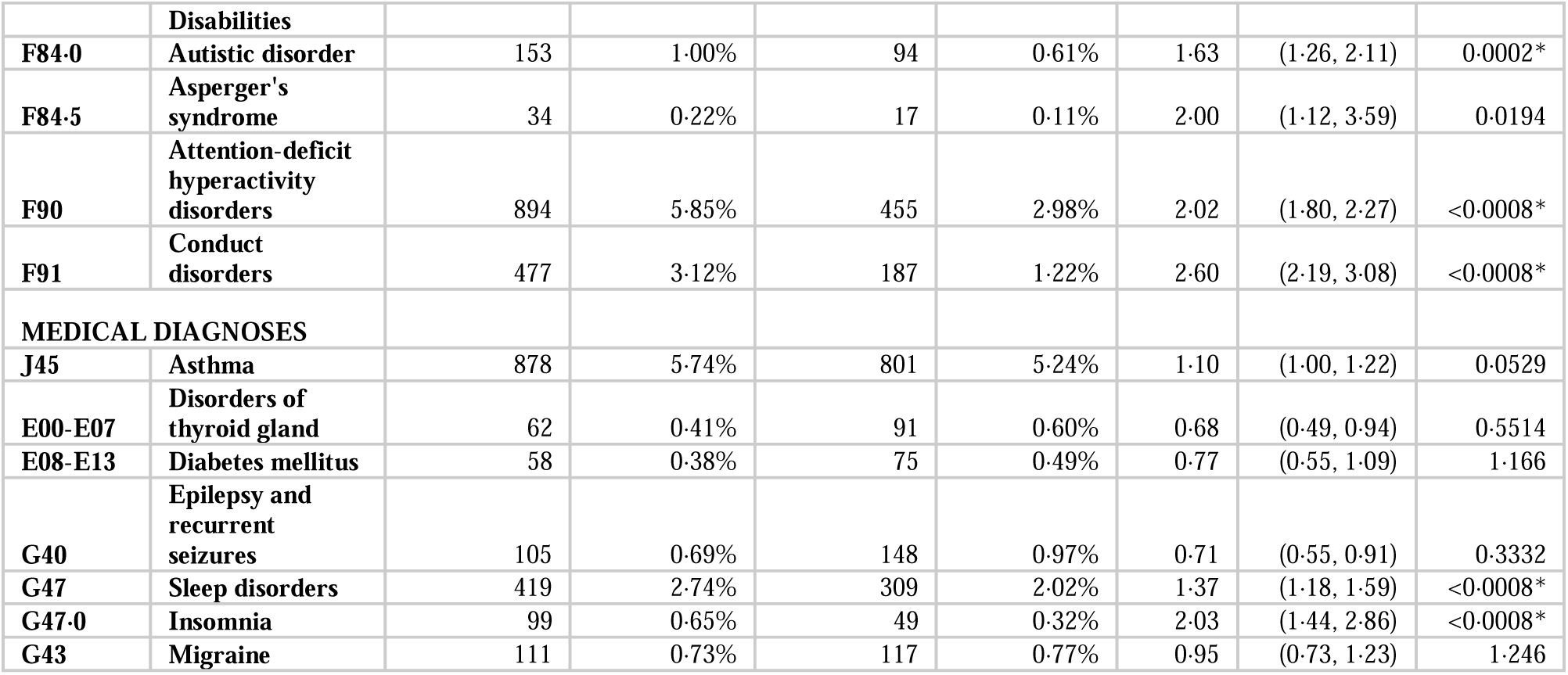
Prevalence and odds ratios of medical/psychiatric/socioeconomic characteristics in patients 13-21 with or without SOSH among COVID-19 (-) patients. Statistical comparisons are represented by odds ratios (OR) with 95% confidence intervals (CI). Significant p-values are indicated with an asterisk (*).

Of the adolescents negative for COVID-19 with SI/SA, the odds were greater for all pre-existing risk factors, with exceptions: one psychiatric risk factor (intellectual disabilities (OR 1·42, 95%CI (1·06,1·91), p=0·0184)) and four medical risk factors: disorders of thyroid gland (OR 0·89, 95%CI (0·72,1·09), p=0·2568), diabetes mellitus (OR 0.92, 95%CI (0·73,1·16), p=0·4918), epilepsy (OR 0·86, 95%CI (0·71,1·04), p=0·1153), p=0·1153), and migraine (OR 1·22, 95%CI (1·06,1·41), p=0·0058) (Table 5) (Figure 4).

**Figure 4.**
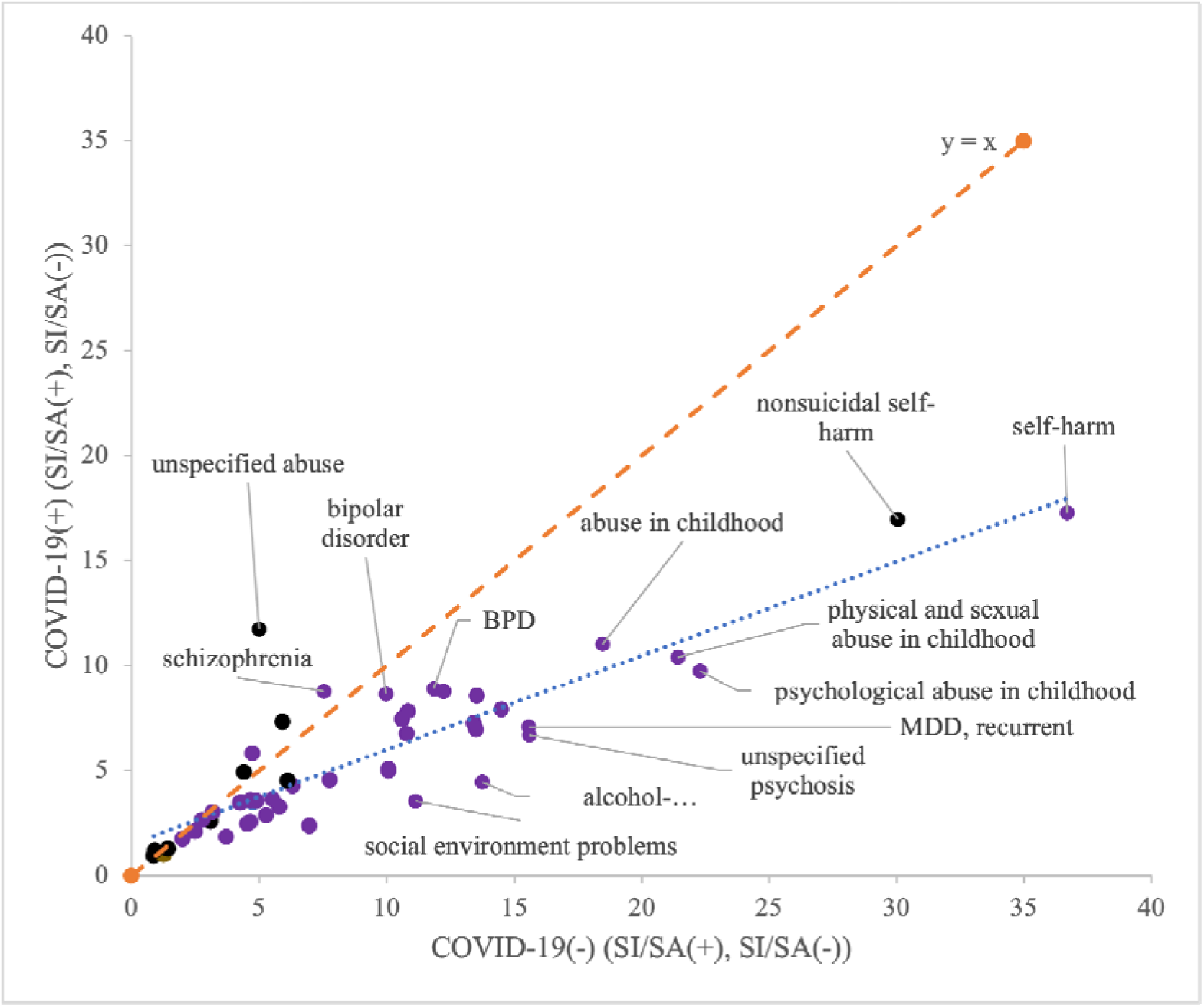
Comparison of odds ratios of medical/psychiatric/socioeconomic characteristics in patients 13-21 years old with or without suicide attempt/suicidal ideation in COVID-19 (-) and COVID-19 (+) patients. Labeled characteristics are a sample of the significant findings. Key: Purple denotes significance in both COVID-19(+) an COVID-19(-) patients; yellow denotes significance in only COVID-19(+) patients; brown denotes significance i only COVID-19(-) patients; and black denotes a lack of significance in each group.

**Table 5.** Prevalence and odds ratios of medical/psychiatric/socioeconomic characteristics in patients 0-12 with or without SI/SA among COVID-19 (-) patients. Statistical comparisons are represented by odds ratios (OR) with 95% confidence intervals (CI). Significant p-values are indicated with an asterisk (*).

| **ICD-10 Code** | **Characteristic** | **COVID-19(-), SI/SA(+)** |  | **COVID-19(-), SI/SA(-)** |  | **OR** | **95%CI** | **P value** |
| --- | --- | --- | --- | --- | --- | --- | --- | --- |
|  |  | **N** | **%** | **N** | **%** |  |  |  |
| **SOCIOECONOMIC DIAGNOSES** |  |  |  |  |  |  |  |  |
| **Z55** | **Problems related to education and literacy** | 344 | 2.22% | 94 | 0.61% | 3.72 | (2.96, 4.68) | <0.0008* |
| **Z59** | **Problems related to housing and economic circumstances** | 99 | 0.64% | 36 | 0.23% | 2.76 | (1.88, 4.05) | <0.0008* |
| **Z60** | **Problems related to social environment** | 209 | 1.35% | 19 | 0.12% | 11.14 | (6.96, 17.82) | <0.0008* |
| **Z62** | **Problems related to upbringing** | 1147 | 7.41% | 114 | 0.74% | 10.79 | (8.89, 13.09) | <0.0008* |
| **Z62-2** | **Upbringing away from parents** | 213 | 1.38% | 50 | 0.32% | 4.31 | (3.16, 5.86) | <0.0008* |
| **Z62.81** | **Personal history of abuse in childhood** | 639 | 4.13% | 36 | 0.23% | 18.47 | (13.19, 25.86) | <0.0008* |
| **Z62.810** | **Personal history of physical and sexual abuse in childhood** | 478 | 3.09% | 23 | 0.15% | 21.41 | (14.08, 32.56) | <0.0008* |
| **Z62-82** | **Parent-child conflict** | 447 | 2.89% | 34 | 0.22% | 13.51 | (9.52, 19.16) | <0.0008* |
| **Z63** | **Other problems related to primary support** group, including family circumstances | 687 | 4.44% | 92 | 0.59% | 7.77 | (6.24, 9.67) | <0.0008* |
| **Z65** | Problems related to other psychosocial circumstances | 275 | 1.78% | 40 | 0.26% | 6.98 | (5.01, 9.73) | <0.0008* |
| **Z81** | Family history of mental and behavioral disorders | 958 | 6.19% | 83 | 0.54% | 12.24 | (9.77, 15.33) | <0.0008* |
| **Z91.5** | Personal history of self-harm | 1674 | 10.82% | 51 | 0.33% | 36.68 | (27.74, 48.52) | <0.0008* |
| **PSYCHIATRIC DIAGNOSES** |  |  |  |  |  |  |  |  |
| **F10-F19** | Mental and behavioral disorders due to psychoactive substance use | 1208 | 7.81% | 129 | 0.83% | 10.07 | (8.39, 12.10) | <0.0008* |
| **F10** | Alcohol related disorders | 244 | 1.58% | 18 | 0.12% | 13.76 | (8.52, 22.21) | <0.0008* |
| **F12** | Cannabis related disorders | 708 | 4.58% | 68 | 0.44% | 10.86 | (8.46, 13.95) | <0.0008* |
| **F17** | Nicotine dependence | 403 | 2.60% | 41 | 0.26% | 10.07 | (7.29, 13.89) | <0.0008* |
| **F20-F29** | Schizophrenia, schizotypal, delusional, and other non-mood psychotic disorders | 319 | 2.06% | 24 | 0.16% | 13.55 | (8.94, 20.53) | <0.0008* |
| **F29** | Unspecified psychosis not due to a substance or known physiological condition | 185 | 1.20% | 12 | 0.08% | 15.59 | (8.69, 27.97) | <0.0008* |
| **F30-F39** | Mood [affective] disorders | 5929 | 38.32% | 636 | 4.11% | 14.49 | (13.30, 15.79) | <0.0008* |
| **F31** | Bipolar disorder | 361 | 2.33% | 37 | 0.24% | 9.97 | (7.10, 13.99) | <0.0008* |
| **F32** | Depressive episode | 4616 | 29.83% | 475 | 3.07% | 13.42 | (12.18, 14.80) | <0.0008* |
| **F33** | Major depressive disorder, recurrent | 1627 | 10.52% | 116 | 0.75% | 15.56 | (12.87, 18.81) | <0.0008* |
| **F34.1** | Dysthymic disorder | 126 | 0.81% | 28 | 0.18% | 4.53 | (3.01, 6.82) | <0.0008* |
| **F40-F48** | Anxiety, dissociative, stress-related, somatoform and other nonpsychotic mental disorders | 4935 | 31.89% | 1206 | 7.79% | 5.54 | (5.18, 5.93) | <0.0008* |
| **F41.0** | Panic disorder [episodic paroxysmal anxiety] | 353 | 2.28% | 57 | 0.37% | 6.31 | (4.77, 8.36) | <0.0008* |
| **F41.1** | Generalized anxiety disorder | 1220 | 7.88% | 247 | 1.60% | 5.28 | (4.59, 6.06) | <0.0008* |
| **F41.9** | Anxiety disorder, unspecified | 2656 | 17.17% | 720 | 4.65% | 4.25 | (3.90, 4.63) | <0.0008* |
| **F42** | Obsessive-compulsive disorder | 257 | 1.66% | 45 | 0.29% | 5.79 | (4.22, 7.95) | <0.0008* |
| **F43.1** | Post-traumatic stress disorder (PTSD) | 915 | 5.91% | 91 | 0.59% | 10.62 | (8.55, 13.19) | <0.0008* |
| F43·2 | Adjustment disorders | 981 | 6·34% | 221 | 1·43% | 4·67 | (4·03, 5·42) | <0·0008* |
| F60-F69 | Disorders of adult personality and behavior | 839 | 5·42% | 188 | 1·21% | 4·66 | (3·97, 5·47) | <0·0008* |
| F60·3 | Borderline personality disorder | 176 | 1·14% | 15 | 0·10% | 11·86 | (7·00, 20·10) | <0·0008* |
| F64 | Gender identity disorders | 243 | 1·57% | 52 | 0·34% | 4·73 | (3·50, 6·39) | <0·0008* |
| F70-F79 | Intellectual Disabilities | 108 | 0·70% | 76 | 0·49% | 1·42 | (1·06, 1·91) | 0·0184 |
| F84·0 | Autistic disorder | 446 | 2·88% | 182 | 1·18% | 2·49 | (2·10, 2·97) | <0·0008* |
| F84·5 | Asperger's syndrome | 60 | 0·39% | 19 | 0·12% | 3·17 | (1·89, 5·31) | <0·0008* |
| F90 | Attention-deficit hyperactivity disorders | 2138 | 13·82% | 760 | 4·91% | 3·10 | (2·85, 3·38) | <0·0008* |
| F91 | Conduct disorders | 1039 | 6·71% | 230 | 1·49% | 4·77 | (4·13, 5·51) | <0·0008* |
| MEDICAL DIAGNOSES |  |  |  |  |  |  |  |  |
| J45 | Asthma | 1570 | 10·15% | 1246 | 8·05% | 1·29 | (1·19, 1·39) | <0·0008* |
| E00-E07 | Disorders of thyroid gland | 177 | 1·14% | 199 | 1·29% | 0·89 | (0·72, 1·09) | 0·2568 |
| E08-E13 | Diabetes mellitus | 141 | 0·91% | 153 | 0·99% | 0·92 | (0·73, 1·16) | 0·4918 |
| G40 | Epilepsy and recurrent seizures | 193 | 1·25% | 225 | 1·45% | 0·86 | (0·71, 1·04) | 0·1153 |
| G47 | Sleep disorders | 1140 | 7·37% | 592 | 3·83% | 2·00 | (1·81, 2·21) | <0·0008* |
| G47·0 | Insomnia | 557 | 3·60% | 117 | 0·76% | 4·90 | (4·01, 5·99) | <0·0008* |
| G43 | Migraine | 425 | 2·75% | 349 | 2·26% | 1·22 | (1·06, 1·41) | 0·0058 |

##### Children v. Adolescents

In the COVID-19(-) populations with SOSH (Table 6) and with SI/SA (Table 7), no characteristics had significantly greater odds in children only. There were greater odds for several characteristics only in adolescents, and for several characteristics in both children in adolescents (Table 6) (Table 7).

**Table 6.** Odds ratios (OR) of medical, psychiatric, and socioeconomic characteristics in COVID-19 (-) patients with or without SOSH, with a comparison between OR for the 0-12 and 13-21 age groups. Bold denotes significantly greater odds. Italics denotes significantly lower odds. Statistical comparisons are represented by odds ratios (OR) with 95% confidence intervals (CI). Significant p-values are indicated with an asterisk. Characteristics observed in a total of 10 or fewer patients were excluded from comparison and are not included in this table.

| ICD-10 Code | Characteristic | 0-12 |  |  | 13-21 |  |  |
| --- | --- | --- | --- | --- | --- | --- | --- |
| SOCIOECONOMIC DIAGNOSES |  | OR | 95%CI | p value | OR | 95%CI | p value |
| Z55 | Problems related to education and literacy |  |  |  | 2·20 | (1·52, 3·19) | <0·0008* |
| Z60 | Problems related to social environment |  |  |  | 2·29 | (1·22, 4·29) | 0·0098 |
| Z62 | Problems related to upbringing | 8·08 | (4·29, 15·22) | *<0·0008 | 3·04 | (2·15, 4·30) | <0·0008* |
| Z62·2 | Upbringing away from parents |  |  |  | 3·13 | (1·78, 5·50) | <0·0008* |
| Z62·82 | Parent-child conflict |  |  |  | 2·54 | (1·34, 4·83) | 0·0045 |
| Z63 | Other problems related to primary support group, including family circumstances |  |  |  | 2·15 | (1·53, 3·01) | <0·0008* |
| Z65 | Problems related to other psychosocial circumstances |  |  |  | 2·45 | (1·41, 4·24) | 0·0014 |
| Z81 | Family history of mental and behavioral disorders |  |  |  | 4·71 | (2·97, 7·46) | <0·0008* |
| **PSYCHIATRIC DIAGNOSES** |  |  |  |  |  |  |  |
| F30-F39 | Mood [affective] disorders |  |  |  | 4·07 | (3·37, 4·92) | <0·0008* |
| F31 | Bipolar disorder |  |  |  | 4·01 | (2·23, 7·21) | <0·0008* |
| F32 | Depressive episode |  |  |  | 3·75 | (2·98, 4·71) | <0·0008* |
| F33 | Major depressive disorder, recurrent |  |  |  | 7·62 | (4·17, 13·92) | <0·0008* |
| F40-F48 | Anxiety, dissociative, stress-related, somatoform and other nonpsychotic mental disorders | 6·82 | (4·94, 9·43) | *<0·0008 | 2·29 | (2·02, 2·60) | <0·0008* |
| F41·0 | Panic disorder [episodic paroxysmal anxiety] |  |  |  | 2·50 | (1·28, 4·89) | 0·0073 |
| F41·1 | Generalized anxiety disorder |  |  |  | 2·27 | (1·75, 2·96) | <0·0008* |
| F41·9 | Anxiety disorder, unspecified | 8·43 | (5·06, 14·05) | *<0·0008 | 2·44 | (2·05, 2·89) | <0·0008* |
| F42 | Obsessive-compulsive disorder |  |  |  | 1·61 | (1·00, 2·58) | 0·0481 |
| F43·1 | Post-traumatic stress disorder (PTSD) |  |  |  | 6·27 | (3·76, 10·47) | <0·0008* |
| F43·2 | Adjustment disorders |  |  |  | 1·90 | (1·50, 2·42) | <0·0008* |
| F60-F69 | Disorders of adult personality and behavior |  |  |  | 2·49 | (2·00, 3·11) | <0·0008* |
| F60·3 | Borderline personality disorder |  |  |  | 4·09 | (2·23, 7·50) | <0·0008* |
| F70-F79 | Intellectual Disabilities |  |  |  | 0·93 | (0·61, 1·42) | 1·207 |
| F84·0 | Autistic disorder | 3·83 | (2·30, 6·40) | *<0·0008 | 1·63 | (1·26, 2·11) | 0·0002* |
| F84·5 | Asperger's syndrome |  |  |  | 2·00 | (1·12, 3·59) | 0·0194 |
| F90 | Attention-deficit hyperactivity disorders | 6·71 | (4·87, 9·24) | *<0·0008 | 2·02 | (1·80, 2·27) | <0·0008* |
| F91 | Conduct disorders | 7·68 | (4·83, 12·21) | *<0·0008 | 2·60 | (2·19, 3·08) | <0·0008* |
| **MEDICAL DIAGNOSES** |  |  |  |  |  |  |  |
| J45 | Asthma | 1·38 | (1·07, 1·78) | 0·0138 | 1·10 | (1·00, 1·22) | 0·0529 |
| E00-E07 | Disorders of thyroid gland |  |  |  | 0·68 | (0·49, 0·94) | 0·5514 |
| E08-E13 | Diabetes mellitus |  |  |  | 0·77 | (0·55, 1·09) | 1·166 |
| **G40** | **Epilepsy and recurrent seizures** | 0.86 | (0.46, 1.60) | 0.6517 | 0.71 | (0.55, 0.91) | 0.3332 |
| **G47** | **Sleep disorders** | **2.01** | **(1.45, 2.80)** | **\*<0.0008** | **1.37** | **(1.18, 1.59)** | **<0.0008\*** |
| **G47.0** | **Insomnia** |  |  |  | **2.03** | **(1.44, 2.86)** | **<0.0008\*** |
| **G43** | **Migraine** | 1.00 | (0.48, 2.11) | 0.9986 | 0.95 | (0.73, 1.23) | 1.246 |

**Table 7.** Odds ratios (OR) of medical, psychiatric, and socioeconomic characteristics in COVID-19 (-) patients with or without SI/SA, with a comparison between OR for the 0-12 and 13-21 age groups. Bold denotes significantly greater odds. Italics denotes significantly lower odds. Statistical comparisons are represented by odds ratios (OR) with 95% confidence intervals (CI). Significant p-values are indicated with an asterisk. Characteristics observed in a total of 10 or fewer patients were excluded from comparison and are not included in this table.

| **ICD-10 Code** | **Characteristic** | **0-12** |  |  | **13-21** |  |  |
| --- | --- | --- | --- | --- | --- | --- | --- |
| **SOCIOECONOMIC DIAGNOSES** |  | **OR** | **95%CI** | **p value** | **OR** | **95%CI** | **p value** |
| **Z55** | Problems related to education and literacy |  |  |  | **3.72** | **(2.96, 4.68)** | **<0.0008\*** |
| **Z59** | Problems related to housing and economic circumstances |  |  |  | **2.76** | **(1.88, 4.05)** | **<0.0008\*** |
| **Z60** | Problems related to social environment |  |  |  | **11.14** | **(6.96, 17.82)** | **<0.0008\*** |
| **Z62** | Problems related to upbringing |  |  |  | **10.79** | **(8.89, 13.09)** | **<0.0008\*** |
| **Z62.2** | Upbringing away from parents |  |  |  | **4.31** | **(3.16, 5.86)** | **<0.0008\*** |
| **Z62.81** | Personal history of abuse in childhood |  |  |  | **18.47** | **(13.19, 25.86)** | **<0.0008\*** |
| **Z62.810** | Personal history of physical and sexual abuse in childhood |  |  |  | **21.41** | **(14.08, 32.56)** | **<0.0008\*** |
| **Z62.82** | Parent-child conflict |  |  |  | **13.51** | **(9.52, 19.16)** | **<0.0008\*** |
| **Z63** | Other problems related to primary support group, including family circumstances |  |  |  | **7.77** | **(6.24, 9.67)** | **<0.0008\*** |
| **Z65** | Problems related to other psychosocial circumstances |  |  |  | **6.98** | **(5.01, 9.73)** | **<0.0008\*** |
| **Z81** | Family history of mental and behavioral disorders |  |  |  | **12.24** | **(9.77, 15.33)** | **<0.0008\*** |
| Z91.5 | Personal history of self-harm |  |  |  | 36.68 | (27.74, 48.52) | <0.0008* |
| PSYCHIATRIC DIAGNOSES |  |  |  |  |  |  |  |
| F10-F19 | Mental and behavioral disorders due to psychoactive substance use |  |  |  | 10.07 | (8.39, 12.10) | <0.0008* |
| F10 | Alcohol related disorders |  |  |  | 13.76 | (8.52, 22.21) | <0.0008* |
| F12 | Cannabis related disorders |  |  |  | 10.86 | (8.46, 13.95) | <0.0008* |
| F17 | Nicotine dependence |  |  |  | 10.07 | (7.29, 13.89) | <0.0008* |
| F20-F29 | Schizophrenia, schizotypal, delusional, and other non-mood psychotic disorders |  |  |  | 13.55 | (8.94, 20.53) | <0.0008* |
| F29 | Unspecified psychosis not due to a substance or known physiological condition |  |  |  | 15.59 | (8.69, 27.97) | <0.0008* |
| F30-F39 | Mood [affective] disorders |  |  |  | 14.49 | (13.30, 15.79) | <0.0008* |
| F31 | Bipolar disorder |  |  |  | 9.97 | (7.10, 13.99) | <0.0008* |
| F32 | Depressive episode |  |  |  | 13.42 | (12.18, 14.80) | <0.0008* |
| F33 | Major depressive disorder, recurrent |  |  |  | 15.56 | (12.87, 18.81) | <0.0008* |
| F34.1 | Dysthymic disorder |  |  |  | 4.53 | (3.01, 6.82) | <0.0008* |
| F40-F48 | Anxiety, dissociative, stress-related, somatoform and other nonpsychotic mental disorders | 8.61 | (6.02, 12.32) | *<0.0008 | 5.54 | (5.18, 5.93) | <0.0008* |
| F41.0 | Panic disorder [episodic paroxysmal anxiety] |  |  |  | 6.31 | (4.77, 8.36) | <0.0008* |
| F41.1 | Generalized anxiety disorder |  |  |  | 5.28 | (4.59, 6.06) | <0.0008* |
| F41.9 | Anxiety disorder, unspecified | 9.44 | (5.50, 16.21) | *<0.0008 | 4.25 | (3.90, 4.63) | <0.0008* |
| F42 | Obsessive-compulsive disorder |  |  |  | 5.79 | (4.22, 7.95) | <0.0008* |
| F43.1 | Post-traumatic stress disorder (PTSD) |  |  |  | 10.62 | (8.55, 13.19) | <0.0008* |
| F43.2 | Adjustment disorders | 6.77 | (3.57, 12.85) | *<0.0008 | 4.67 | (4.03, 5.42) | <0.0008* |
| F60-F69 | Disorders of adult personality and behavior |  |  |  | 4.66 | (3.97, 5.47) | <0.0008* |
| F60.3 | Borderline personality disorder |  |  |  | 11.86 | (7.00, 20.10) | <0.0008* |
| F64 | Gender identity disorders |  |  |  | 4.73 | (3.50, 6.39) | <0.0008* |
| **F70-F79** | **Intellectual Disabilities** |  |  |  | **1.42** | **(1.06, 1.91)** | **0.0184** |
| **F84.0** | **Autistic disorder** | **4.30** | **(2.47, 7.46)** | **\*<0.0008** | 2.49 | (2.10, 2.97) | <0.0008* |
| **F84.5** | **Asperger's syndrome** |  |  |  | **3.17** | **(1.89, 5.31)** | **<0.0008\*** |
| **F90** | **Attention-deficit hyperactivity disorders** | **9.04** | **(6.27, 13.06)** | **\*<0.0008** | **3.10** | **(2.85, 3.38)** | **<0.0008\*** |
| **F91** | **Conduct disorders** | **10.82** | **(6.53, 17.95)** | **\*<0.0008** | **4.77** | **(4.13, 5.51)** | **<0.0008\*** |
| **MEDICAL DIAGNOSES** |  |  |  |  |  |  |  |
| **J45** | **Asthma** | 1.07 | (0.82, 1.40) | 0.6419 | **1.29** | **(1.19, 1.39)** | **<0.0008\*** |
| **E00-E07** | **Disorders of thyroid gland** |  |  |  | 0.89 | (0.72, 1.09) | 0.2568 |
| **E08-E13** | **Diabetes mellitus** |  |  |  | **0.92** | **(0.73, 1.16)** | **0.4918** |
| **G40** | **Epilepsy and recurrent seizures** | 0.70 | (0.33, 1.48) | 0.3598 | **0.86** | **(0.71, 1.04)** | **0.1153** |
| **G47** | **Sleep disorders** | **2.00** | **(1.41, 2.83)** | **\*0.0001** | **2.00** | **(1.81, 2.21)** | **<0.0008\*** |
| **G47.0** | **Insomnia** |  |  |  | **4.90** | **(4.01, 5.99)** | **<0.0008\*** |
| **G43** | **Migraine** |  |  |  | 1.22 | (1.06, 1.41) | 0.0058 |

### Association with Suicidality and Self-Harm in COVID-19(+) Patients

#### SOSH and SI/SA

##### Ages 0-12

Among patients diagnosed with COVID-19, those with later SOSH had a greater odds of psychiatric diagnoses (ADHD (OR 5·56, 95%CI (4·19, 7·38), p<0·0008), conduct disorders (OR 7·32, 95%CI (4·77, 11·24), p<0·0008), adult personality disorders (OR 4·47, 95%CI (2·37, 8·40), p<0·0008), unspecified anxiety (OR 6·18, 95%CI (4·58, 8·34), p<0·0008), generalized anxiety (OR 5·89, 95%CI (3·85, 9·01), p<0·0008)), socioeconomic factors (problems with primary support group (OR 7·06, 95%CI (3·73, 13·36), p<0·0008)), medical diagnoses (thyroid disorders (OR 1·63, 95%CI (1·31, 2·02), p<0·0008), and insomnia (OR 2·46, 95%CI (1·81, 3·34), p<0·0008)) (Table 8) (Figure 1).

**Table 8.** Prevalence and odds ratios of medical/psychiatric/socioeconomic characteristics in patients 0-12 with or without SOSH among COVID-19 (+) patients. Statistical comparisons are represented by odds ratios (OR) with 95% confidence intervals (CI). Significant p-values are indicated with an asterisk (*).

| **ICD-10 Code** | **Characteristic** | **COVID-19(+), SOSH(+)** |  | **COVID-19(+), SOSH(-)** |  | **OR** | **95%CI** | **P value** |
| --- | --- | --- | --- | --- | --- | --- | --- | --- |
|  |  | **N** | **%** | **N** | **%** |  |  |  |
| **SOCIOECONOMIC DIAGNOSES** |  |  |  |  |  |  |  |  |
| **Z59** | **Problems related to housing and economic circumstances** | 15 | 1.10% | 11 | 0.81% | 1.37 | (0.63, 2.99) | 0.4407 |
| **Z63** | **Other problems related to primary support group, including family circumstances** | 74 | 5.45% | 11 | 0.81% | 7.06 | (3.73, 13.36) | *<0.0008 |
| **PSYCHIATRIC DIAGNOSES** |  |  |  |  |  |  |  |  |
| **F41.1** | **Generalized anxiety disorder** | 140 | 10.31% | 26 | 1.91% | 5.89 | (3.85, 9.01) | *<0.0008 |
| **F41.9** | **Anxiety disorder, unspecified** | 281 | 20.69% | 55 | 4.05% | 6.18 | (4.58, 8.34) | *<0.0008 |
| **F60-F69** | **Disorders of adult personality and behavior** | 52 | 3.83% | 12 | 0.88% | 4.47 | (2.37, 8.40) | *<0.0008 |
| **F84.5** | **Asperger's syndrome** | 47 | 3.46% | 31 | 2.28% | 1.53 | (0.97, 2.43) | 0.0675 |
| **F90** | **Attention-deficit hyperactivity disorders** | 293 | 21.58% | 64 | 4.71% | 5.56 | (4.19, 7.38) | *<0.0008 |
| **F91** | **Conduct disorders** | 164 | 12.08% | 25 | 1.84% | 7.32 | (4.77, 11.24) | *<0.0008 |
| **MEDICAL DIAGNOSES** |  |  |  |  |  |  |  |  |
| **E00-E07** | **Disorders of thyroid gland** | 244 | 17.97% | 161 | 11.86% | 1.63 | (1.31, 2.02) | *<0.0008 |
| **G47** | **Sleep disorders** | 29 | 2.14% | 13 | 0.96% | 2.26 | (1.17, 4.36) | 0.0153 |
| **G47.0** | **Insomnia** | 145 | 10.68% | 63 | 4.64% | 2.46 | (1.81, 3.34) | *<0.0008 |

Patients with SI/SA also had greater odds of psychiatric diagnoses (ADHD (OR 7·93, 95%CI (5·75, 10·93), p<0·0008), conduct disorders (OR 11·50, 95%CI (6·84, 19·34), p<0·0008), adjustment disorders (OR 7·04, 95%CI (3·82, 12·97), p<0·0008), personality disorders (OR 3·67, 95%CI (2·02, 6·68), p<0·0008), anxiety disorders (OR 5·69, 95%CI (4·23, 7.·64), p<0·0008), unspecified anxiety (OR 5·03, 95%CI (3·36, 7·52), p<0·0008)), socioeconomic factors (problems related to primary support group (OR 5·69, 95%CI (3·26, 9·93), p<0·0008)), and medical diagnoses (asthma (OR 1·59, 95%CI (1·28, 1·98), p<0·0008), sleep disorders (OR 2·22, 95%CI (1·64, 3·00), p<0·0008)) (Table 9) (Figure 2).

**Table 9.** Prevalence and odds ratios of medical/psychiatric/socioeconomic characteristics in patients 0-12 with or without SI/SA among COVID-19 (+) patients. Statistical comparisons are represented by odds ratios (OR) with 95% confidence intervals (CI). Significant p-values are indicated with an asterisk (*).

| **ICD-10 Code** | **Characteristic** | **COVID-19(+), SI/SA(+)** |  | **COVID-19(+), SI/SA(-)** |  | **OR** | **95%CI** | **P value** |
| --- | --- | --- | --- | --- | --- | --- | --- | --- |
|  |  | **N** | **%** | **N** | **%** |  |  |  |
| **SOCIOECONOMIC DIAGNOSES** |  |  |  |  |  |  |  |  |
| **Z63** | **Other problems related to primary support group, including family circumstances** | 81 | 6.21% | 15 | 1.15% | 5.69 | (3.26, 9.93) | *<0.0008 |
| **PSYCHIATRIC DIAGNOSES** |  |  |  |  |  |  |  |  |
| **F40-F48** | **Anxiety, dissociative, stress-related, somatoform and other nonpsychotic mental disorders** | 273 | 20.94% | 58 | 4.45% | 5.69 | (4.23, 7.64) | *<0.0008 |
| **F41.9** | **Anxiety disorder, unspecified** | 138 | 10.58% | 30 | 2.30% | 5.03 | (3.36, 7.52) | *<0.0008 |
| **F43.2** | **Adjustment disorders** | 80 | 6.13% | 12 | 0.92% | 7.04 | (3.82, 12.97) | *<0.0008 |
| **F60-F69** | **Disorders of adult personality and behavior** | 50 | 3.83% | 14 | 1.07% | 3.67 | (2.02, 6.68) | *<0.0008 |
| **F84.0** | **Autistic disorder** | 42 | 3.22% | 16 | 1.23% | 2.68 | (1.50, 4.79) | 0.0009 |
| **F90** | **Attention-deficit hyperactivity disorders** | 293 | 22.47% | 46 | 3.53% | 7.93 | (5.75, 10.93) | *<0.0008 |
| **F91** | **Conduct disorders** | 163 | 12.50% | 16 | 1.23% | 11.50 | (6.84, 19.34) | *<0.0008 |
| **MEDICAL DIAGNOSES** |  |  |  |  |  |  |  |  |
| **J45** | **Asthma** | 229 | 17.56% | 154 | 11.81% | 1.59 | (1.28, 1.98) | *<0.0008 |
| **G40** | **Epilepsy and recurrent seizures** | 24 | 1.84% | 16 | 1.23% | 1.51 | (0.80, 2.85) | 0.20701 |
| **G47** | **Sleep disorders** | 140 | 10.74% | 67 | 5.14% | 2.22 | (1.64, 3.00) | *<0.0008 |
| **G43** | **Migraine** | 12 | 0.92% | 12 | 0.92% | 1.00 | (0.45, 2.23) | 1 |

##### Ages 13-21

In the adolescent COVID-19-positive group, those with SOSH had greater odds of psychiatric risk factors including ADHD (OR 1·93; 95% CI (1·75, 2·13), p=<0·0008), conduct disorders (OR 2·51; 95% CI (2·17,2·90), p=<0·0008), adjustment disorders (OR 1·73 ; CI (1·43, 2·10), p=<0·0008) , PTSD (OR 3·84 ; 95% CI (2·62,5·64), p=<0·0008), bipolar disorder (OR 6·28 ; 95% CI (3·41, 11·55), p=<0·0008), disorders of adults personality and behavior (OR 1·96 ; 95% CI (1·62, 2·36), p=<0·0008), borderline personality disorder (OR 5·38 ; 95% CI (2·83,10·25), p=<0·0008), mood disorders (OR 3·26 ; 95% CI (2·79, 3·82), p=<0·0008), anxiety and non-psychotic disorders (OR 1·77 ; 95% CI (1·60, 1·96), p=<0·0008), unspecified anxiety disorder (OR 1·70 ; 95% CI (1·49, 1·95), p=<0·0008), generalized anxiety disorder ( OR 1·84 ; 95% CI (1·47, 2·31), p=<0·0008), depressive episode (OR 3·26 ; 95% CI (2·70, 3·94), p=<0·0008), recurrent major depressive disorder (OR 5·38 ; 95% CI (3·20, 9·03), p=<0·0008) and autism (OR 1·67 ; 95% CI (1·31, 2·11), p=<0·0008) (Table 10) (Figure 3). Patients had greater odds of socioeconomic factors including problems related to primary support groups (OR 2·58; 95% CI (1·90,3·50), p=<0·0008), problems related to upbringing (OR 3·75; 95% CI (2·64,5·32), p= <0·0008), family history of mental and behavioral disorders (OR 3·64; 95% CI (2·41,5·49), p=<0·0008), parent-child conflict (OR 2·85; 95% CI (1·69,4·81), p=0·0001) and upbringing away from parents (OR 3·51; 95% CI (1·85, 6·66), p=0·0001). These patients had greater odds of sleep disorders (OR 1·25; 95% CI [1·10, 1·42], p=0·0005) and insomnia (OR 2·15; 95% CI [1·62, 2·85], p=<0·0008).

**Table 10.** Prevalence and odds ratios of medical/psychiatric/socioeconomic characteristics in patients 13-21 with or without SOSH among COVID-19 (+) patients. Statistical comparisons are represented by odds ratios (OR) with 95% confidence intervals (CI). Significant p-values are indicated with an asterisk (*).

| ICD-10 Code | Characteristic | COVID-19(+), SOSH(+) |  | COVID-19(+), SOSH(-) |  | OR | 95% CI | P value |
| --- | --- | --- | --- | --- | --- | --- | --- | --- |
|  |  | N | % | N | % |  |  |  |
| **SOCIOECONOMIC DIAGNOSES** |  |  |  |  |  |  |  |  |
| Z55 | Problems related to education and literacy | 80 | 0.52% | 67 | 0.44% | 1.20 | (0.86, 1.65) | 0.2868 |
| Z60 | Problems related to social environment | 25 | 0.16% | 15 | 0.10% | 1.67 | (0.88, 3.16) | 0.1175 |
| Z62 | Problems related to upbringing | 149 | 0.97% | 40 | 0.26% | 3.75 | (2.64, 5.32) | <0.0008* |
| Z62.2 | Upbringing away from parents | 42 | 0.27% | 12 | 0.08% | 3.51 | (1.85, 6.66) | 0.0001* |
| Z62.82 | Parent-child conflict | 54 | 0.35% | 19 | 0.12% | 2.85 | (1.69, 4.81) | 0.0001* |
| Z63 | Other problems related to primary support group, including family circumstances | 146 | 0.95% | 57 | 0.37% | 2.58 | (1.90, 3.50) | <0.0008* |
| Z65 | Problems related to other psychosocial circumstances | 62 | 0.41% | 40 | 0.26% | 1.55 | (1.04, 2.31) | 0.03016 |
| Z81 | Family history of mental and behavioral disorders | 105 | 0.69% | 29 | 0.19% | 3.64 | (2.41, 5.49) | <0.0008* |
| **PSYCHIATRIC DIAGNOSES** |  |  |  |  |  |  |  |  |
| F10-F19 | Mental and behavioral disorders due to psychoactive substance use | 49 | 0.32% | 29 | 0.19% | 1.69 | (1.07, 2.68) | 0.0248 |
| F30-F39 | Mood [affective] disorders | 668 | 4.37% | 211 | 1.38% | 3.26 | (2.79, 3.82) | <0.0008* |
| F31 | Bipolar disorder | 75 | 0.49% | 12 | 0.08% | 6.28 | (3.41, 11.55) | <0.0008* |
| F32 | Depressive episode | 460 | 3.01% | 144 | 0.94% | 3.26 | (2.70, 3.94) | <0.0008* |
| F33 | Major depressive disorder, recurrent | 91 | 0.59% | 17 | 0.11% | 5.38 | (3.20, 9.03) | <0.0008* |
| F34.1 | Dysthymic disorder | 22 | 0.14% | 13 | 0.08% | 1.69 | (0.85, 3.36) | 0.1325 |
| F40-F48 | Anxiety, dissociative, stress-related, somatoform and other nonpsychotic mental disorders | 1066 | 6.97% | 622 | 4.07% | 1.77 | (1.60, 1.96) | <0.0008* |
| F41.0 | Panic disorder [episodic paroxysmal anxiety] | 42 | 0.27% | 19 | 0.12% | 2.21 | (1.29, 3.81) | 0.0041 |
| F41.1 | Generalized **anxiety disorder** | 216 | 1.41% | 118 | 0.77% | 1.84 | (1.47, 2.31) | <0.0008* |
| **F41-9** | **Anxiety disorder, unspecified** | 577 | 3.77% | 344 | 2.25% | 1.70 | (1.49, 1.95) | <0.0008* |
| **F42** | **Obsessive-compulsive disorder** | 59 | 0.39% | 32 | 0.21% | 1.85 | (1.20, 2.84) | 0.0053 |
| **F43-1** | **Post-traumatic stress disorder (PTSD)** | 126 | 0.82% | 33 | 0.22% | 3.84 | (2.62, 5.64) | <0.0008* |
| **F43-2** | **Adjustment disorders** | 280 | 1.83% | 163 | 1.07% | 1.73 | (1.43, 2.10) | <0.0008* |
| **F60-F69** | **Disorders of adult personality and behavior** | 331 | 2.16% | 171 | 1.12% | 1.96 | (1.62, 2.36) | <0.0008* |
| **F60-3** | **Borderline personality disorder** | 59 | 0.39% | 11 | 0.07% | 5.38 | (2.83, 10.25) | <0.0008* |
| **F70-F79** | **Intellectual Disabilities** | 45 | 0.29% | 67 | 0.44% | 0.67 | (0.46, 0.98) | 0.0382 |
| **F84-0** | **Autistic disorder** | 184 | 1.20% | 111 | 0.73% | 1.67 | (1.31, 2.11) | <0.0008* |
| **F84-5** | **Asperger's syndrome** | 40 | 0.26% | 17 | 0.11% | 2.36 | (1.34, 4.16) | 0.0031 |
| **F90** | **Attention-deficit hyperactivity disorders** | 1181 | 7.72% | 636 | 4.16% | 1.93 | (1.75, 2.13) | <0.0008* |
| **F91** | **Conduct disorders** | 633 | 4.14% | 259 | 1.69% | 2.51 | (2.17, 2.90) | <0.0008* |
| **MEDICAL DIAGNOSES** |  |  |  |  |  |  |  |  |
| **J45** | **Asthma** | 1172 | 7.66% | 1285 | 8.40% | 0.90 | (0.83, 0.98) | 0.0174 |
| **E00-E07** | **Disorders of thyroid gland** | 70 | 0.46% | 93 | 0.61% | 0.75 | (0.55, 1.03) | 0.0714 |
| **E08-E13** | **Diabetes mellitus** | 88 | 0.58% | 75 | 0.49% | 1.17 | (0.86, 1.60) | 0.3122 |
| **G40** | **Epilepsy and recurrent seizures** | 130 | 0.85% | 171 | 1.12% | 0.76 | (0.60, 0.95) | 0.0178 |
| **G47** | **Sleep disorders** | 562 | 3.67% | 452 | 2.96% | 1.25 | (1.10, 1.42) | 0.0005* |
| **G47-0** | **Insomnia** | 154 | 1.01% | 72 | 0.47% | 2.15 | (1.62, 2.85) | <0.0008* |
| **G43** | **Migraine** | 200 | 1.31% | 179 | 1.17% | 1.12 | (0.91, 1.37) | 0.2815 |

Among the COVID-19 positive adolescents who developed SI/SA, odds were greater for all pre-existing risk factors with a few exceptions: one psychiatric risk factor (intellectual disabilities (OR 1·28, 95%CI (0·99,1·64), p=0·057)) and five medical risk factors (asthma (OR 1·01, 95%CI (0·94,1·08), p=0·8581), disorders of thyroid gland (OR 1·00, 95%CI (0·82,1·22), p=1·000), diabetes mellitus (OR 1·18, 95%CI (0·97,1·43), p=0·0889), epilepsy (OR 0·94, 95%CI (0·79,1·12), p=0·4811), and migraine (OR 1·01, 95%CI (0·89,1·15), p=0·8796)) (Table 11) (Figure 4).

**Table 11.**
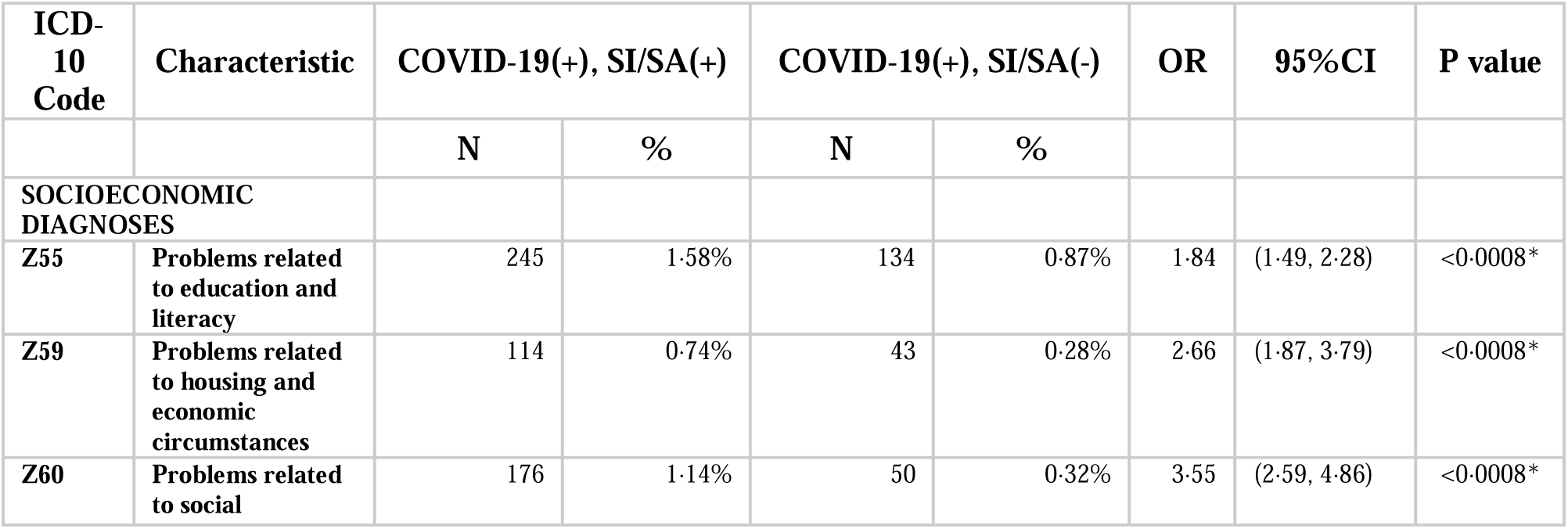
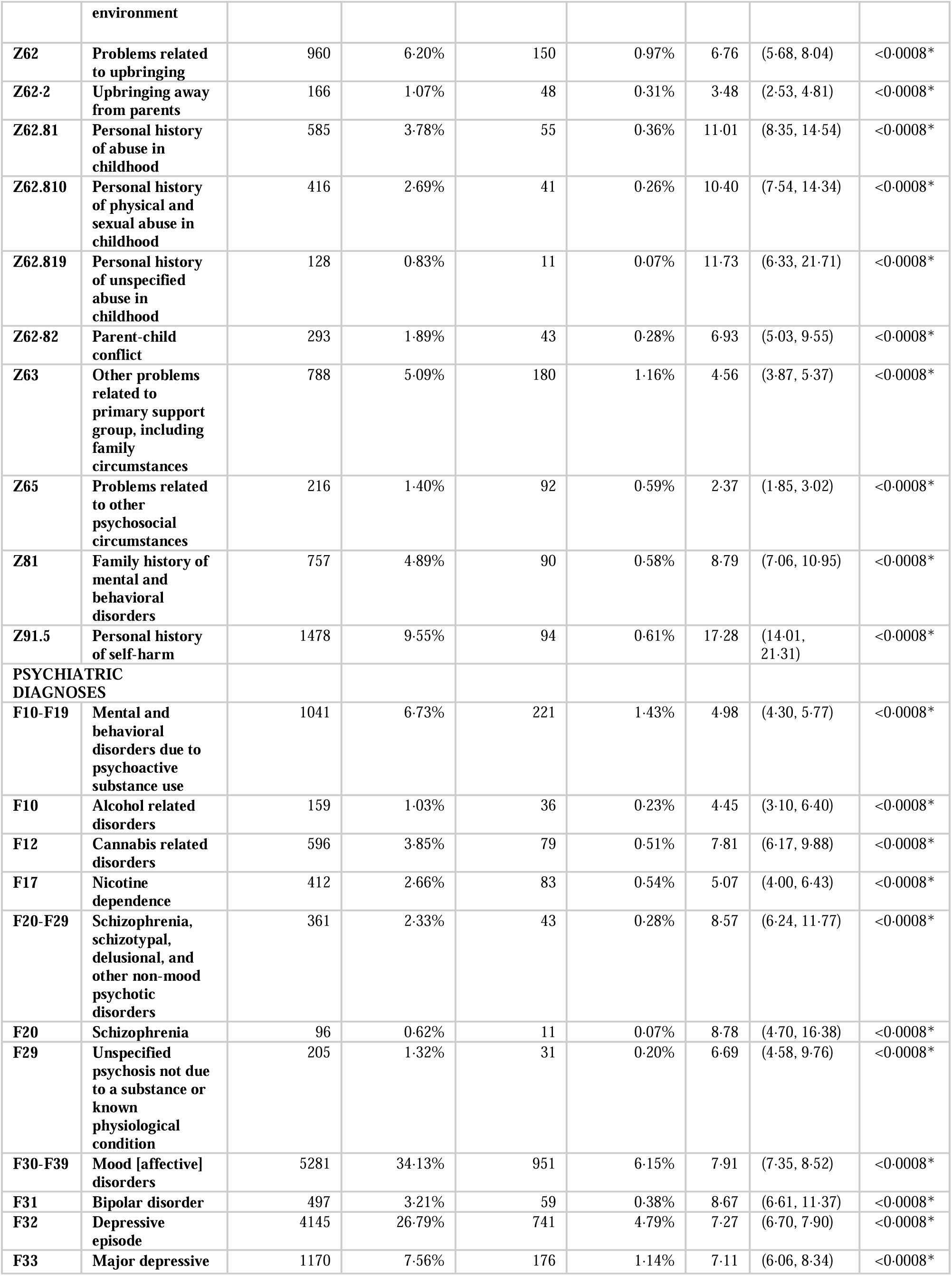
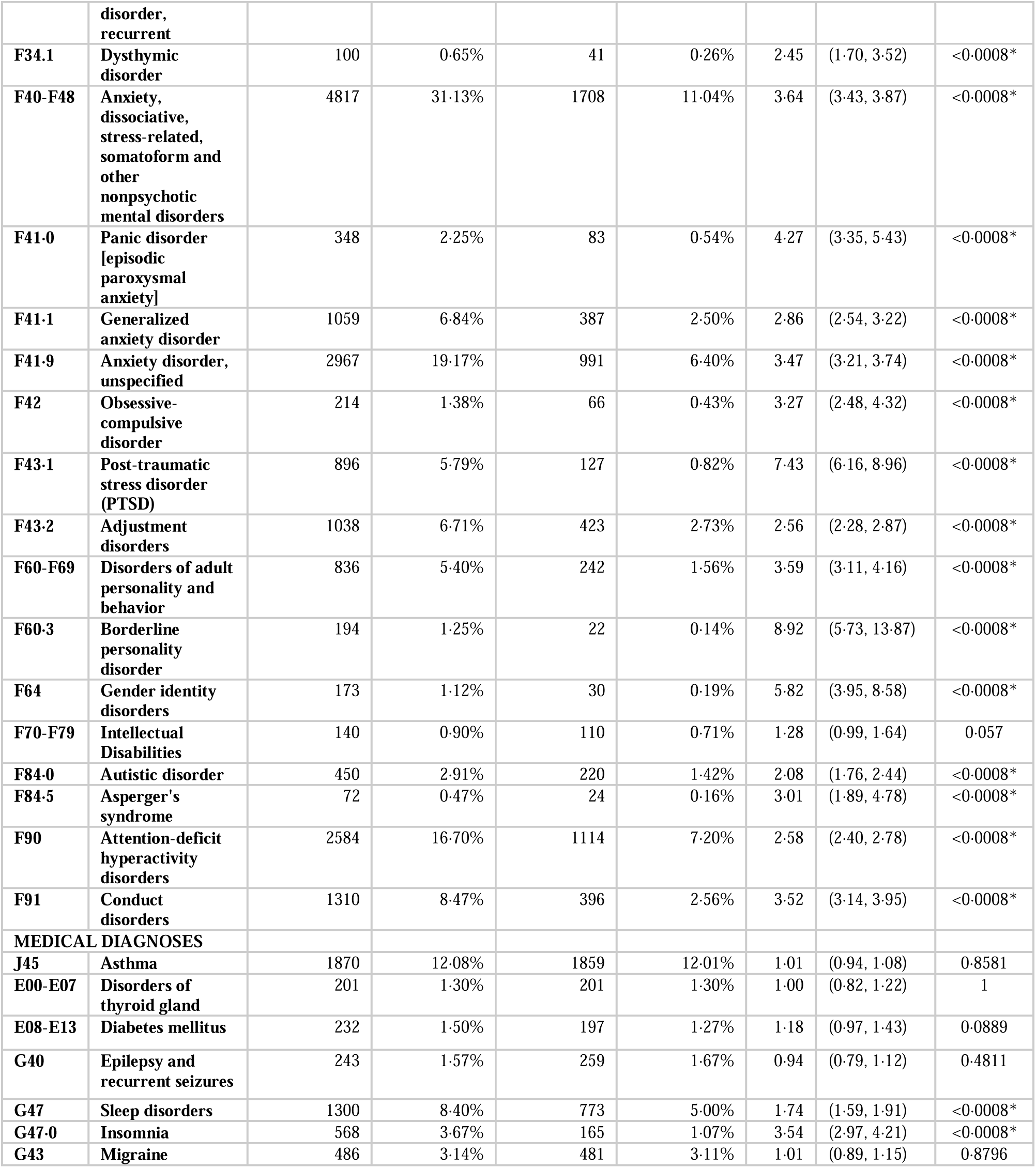
Prevalence and odds ratios of medical/psychiatric/socioeconomic characteristics in patients 13-21 with or without SI/SA among COVID-19 (+) patients. Statistical comparisons are represented by odds ratios (OR) with 95% confidence intervals (CI). Significant p-values are indicated with an asterisk (*).

### Children vs. Adolescents

In the COVID-19(+) populations with SOSH (Table 12), there were significantly greater odds of one characteristic in children only, several in adolescents only, and several in both children and adolescents. In the COVID-19(+) population with SI/SA (Table 13), there were also significantly greater odds of only one characteristic in children only, several in exclusively adolescents, and several in both groups.

**Table 12.** Odds ratios (OR) of medical, psychiatric, and socioeconomic characteristics in COVID-19 (+) patients with or without SOSH, with a comparison between OR for the 0-12 and 13-21 age groups. Bold denotes significantly greater odds. Italics denotes significantly lower odds. Statistical comparisons are represented by odds ratios (OR) with 95% confidence intervals (CI). Significant p-values are indicated with an asterisk. Characteristics observed in a total of 10 or fewer patients were excluded from comparison and are not included in this table.

| ICD-10 Code | Characteristic | 0-12 |  |  | 13-21 |  |  |
| --- | --- | --- | --- | --- | --- | --- | --- |
| SOCIOECONOMIC DIAGNOSES |  | OR | 95% CI | p value | OR | 95% CI | p value |
| Z55 | Problems related to education and literacy |  |  |  | 1.20 | (0.86, 1.65) | 0.2868 |
| Z59 | Problems related to housing and economic circumstances | 1.37 | (0.63, 2.99) | 0.4407 |  |  |  |
| Z60 | Problems related to social environment |  |  |  | 1.67 | (0.88, 3.16) | 0.1175 |
| Z62 | Problems related to upbringing |  |  |  | 3.75 | (2.64, 5.32) | <0.0008* |
| Z62.2 | Upbringing away from parents |  |  |  | 3.51 | (1.85, 6.66) | 0.0001* |
| Z62.82 | Parent-child conflict |  |  |  | 2.85 | (1.69, 4.81) | 0.0001* |
| Z63 | Other problems related to primary support group, including family circumstances | 7.06 | (3.73, 13.36) | *<0.0008 | 2.58 | (1.90, 3.50) | <0.0008* |
| Z65 | Problems related to other psychosocial circumstances |  |  |  | 1.55 | (1.04, 2.31) | 0.03016 |
| Z81 | Family history of mental and behavioral disorders |  |  |  | 3.64 | (2.41, 5.49) | <0.0008* |
| PSYCHIATRIC DIAGNOSES |  |  |  |  |  |  |  |
| F10-F19 | Mental and behavioral disorders due to psychoactive substance use |  |  |  | 1.69 | (1.07, 2.68) | 0.0248 |
| F30-F39 | Mood [affective] disorders |  |  |  | 3.26 | (2.79, 3.82) | <0.0008* |
| F31 | Bipolar disorder |  |  |  | 6.28 | (3.41, 11.55) | <0.0008* |
| F32 | Depressive episode |  |  |  | 3.26 | (2.70, 3.94) | <0.0008* |
| F33 | Major depressive disorder, recurrent |  |  |  | 5.38 | (3.20, 9.03) | <0.0008* |
| F34.1 | Dysthymic disorder |  |  |  | 1.69 | (0.85, 3.36) | 0.1325 |
| F40-F48 | Anxiety, dissociative, stress-related, somatoform and other nonpsychotic mental disorders |  |  |  | 1.77 | (1.60, 1.96) | <0.0008* |
| F41.0 | Panic disorder [episodic paroxysmal anxiety] |  |  |  | 2.21 | (1.29, 3.81) | 0.0041 |
| F41.1 | Generalized anxiety disorder | 5.89 | (3.85, 9.01) | *<0.0008 | 1.84 | (1.47, 2.31) | <0.0008* |
| F41.9 | Anxiety disorder, unspecified | 6.18 | (4.58, 8.34) | *<0.0008 | 1.70 | (1.49, 1.95) | <0.0008* |
| **F42** | **Obsessive-compulsive disorder** |  |  |  | 1.85 | (1.20, 2.84) | 0.0053 |
| **F43.1** | **Post-traumatic stress disorder (PTSD)** |  |  |  | **3.84** | **(2.62, 5.64)** | **<0.0008\*** |
| **F43.2** | **Adjustment disorders** |  |  |  | **1.73** | **(1.43, 2.10)** | **<0.0008\*** |
| **F60-F69** | **Disorders of adult personality and behavior** | **4.47** | **(2.37, 8.40)** | **\*<0.0008** | **1.96** | **(1.62, 2.36)** | **<0.0008\*** |
| **F60.3** | **Borderline personality disorder** |  |  |  | **5.38** | **(2.83, 10.25)** | **<0.0008\*** |
| **F70-F79** | **Intellectual Disabilities** |  |  |  | **0.67** | **(0.46, 0.98)** | **0.0382** |
| **F84.0** | **Autistic disorder** |  |  |  | **1.67** | **(1.31, 2.11)** | **<0.0008\*** |
| **F84.5** | **Asperger's syndrome** | 1.53 | (0.97, 2.43) | 0.0675 | 2.36 | (1.34, 4.16) | 0.0031 |
| **F90** | **Attention-deficit hyperactivity disorders** | **5.56** | **(4.19, 7.38)** | **\*<0.0008** | **1.93** | **(1.75, 2.13)** | **<0.0008\*** |
| **F91** | **Conduct disorders** | **7.32** | **(4.77, 11.24)** | **\*<0.0008** | **2.51** | **(2.17, 2.90)** | **<0.0008\*** |
| **MEDICAL DIAGNOSES** |  |  |  |  |  |  |  |
| **J45** | **Asthma** |  |  |  | 0.90 | (0.83, 0.98) | 0.0174 |
| **E00-E07** | **Disorders of thyroid gland** | **1.63** | **(1.31, 2.02)** | **\*<0.0008** | 0.75 | (0.55, 1.03) | 0.0714 |
| **E08-E13** | **Diabetes mellitus** |  |  |  | 1.17 | (0.86, 1.60) | 0.3122 |
| **G40** | **Epilepsy and recurrent seizures** |  |  |  | 0.76 | (0.60, 0.95) | 0.0178 |
| **G47** | **Sleep disorders** | 2.26 | (1.17, 4.36) | 0.0153 | **1.25** | **(1.10, 1.42)** | **0.0005\*** |
| **G47.0** | **Insomnia** | **2.46** | **(1.81, 3.34)** | **\*<0.0008** | **2.15** | **(1.62, 2.85)** | **<0.0008\*** |
| **G43** | **Migraine** |  |  |  | 1.12 | (0.91, 1.37) | 0.2815 |

**Table 13.** Odds ratios (OR) of medical, psychiatric, and socioeconomic characteristics in COVID-19 (+) patients with or without SI/SA, with a comparison between OR for the 0-12 and 13-21 age groups. Bold denotes significantly greater odds. Italics denotes significantly lower odds. Statistical comparisons are represented by odds ratios (OR) with 95% confidence intervals (CI). Significant p-values are indicated with an asterisk. Characteristics observed in a total of 10 or fewer patients were excluded from comparison and are not included in this table.

| **ICD-10 Code** | **Characteristic** | **0-12** |  |  | **13-21** |  |  |
| --- | --- | --- | --- | --- | --- | --- | --- |
| **SOCIOECONOMIC DIAGNOSES** |  | **OR** | **95%CI** | **p value** | **OR** | **95%CI** | **p value** |
| **Z55** | **Problems related to education and literacy** |  |  |  | **1.84** | **(1.49, 2.28)** | **<0.0008\*** |
| **Z59** | **Problems related to housing and economic circumstances** |  |  |  | **2.66** | **(1.87, 3.79)** | **<0.0008\*** |
| **Z60** | **Problems related to social environment** |  |  |  | **3.55** | **(2.59, 4.86)** | **<0.0008\*** |
| **Z62** | **Problems related** to upbringing |  |  |  | **6.76** | **(5.68, 8.04)** | **<0.0008\*** |
| Z62.2 | Upbringing away from parents |  |  |  | 3.48 | (2.53, 4.81) | <0.0008* |
| Z62.81 | Personal history of abuse in childhood |  |  |  | 11.01 | (8.35, 14.54) | <0.0008* |
| Z62.810 | Personal history of physical and sexual abuse in childhood |  |  |  | 10.40 | (7.54, 14.34) | <0.0008* |
| Z62.819 | Personal history of unspecified abuse in childhood |  |  |  | 11.73 | (6.33, 21.71) | <0.0008* |
| Z62.82 | Parent-child conflict |  |  |  | 6.93 | (5.03, 9.55) | <0.0008* |
| Z63 | Other problems related to primary support group, including family circumstances | 5.69 | (3.26, 9.93) | *<0.0008 | 4.56 | (3.87, 5.37) | <0.0008* |
| Z65 | Problems related to other psychosocial circumstances |  |  |  | 2.37 | (1.85, 3.02) | <0.0008* |
| Z81 | Family history of mental and behavioral disorders |  |  |  | 8.79 | (7.06, 10.95) | <0.0008* |
| Z91.5 | Personal history of self-harm |  |  |  | 17.28 | (14.01, 21.31) | <0.0008* |
| PSYCHIATRIC DIAGNOSES |  |  |  |  |  |  |  |
| F10-F19 | Mental and behavioral disorders due to psychoactive substance use |  |  |  | 4.98 | (4.30, 5.77) | <0.0008* |
| F10 | Alcohol related disorders |  |  |  | 4.45 | (3.10, 6.40) | <0.0008* |
| F12 | Cannabis related disorders |  |  |  | 7.81 | (6.17, 9.88) | <0.0008* |
| F17 | Nicotine dependence |  |  |  | 5.07 | (4.00, 6.43) | <0.0008* |
| F20-F29 | Schizophrenia, schizotypal, delusional, and other non-mood psychotic disorders |  |  |  | 8.57 | (6.24, 11.77) | <0.0008* |
| F20 | Schizophrenia |  |  |  | 8.78 | (4.70, 16.38) | <0.0008* |
| F29 | Unspecified psychosis not due to a substance or known physiological condition |  |  |  | 6.69 | (4.58, 9.76) | <0.0008* |
| F30-F39 | Mood [affective] disorders |  |  |  | 7.91 | (7.35, 8.52) | <0.0008* |
| F31 | Bipolar disorder |  |  |  | 8.67 | (6.61, 11.37) | <0.0008* |
| F32 | Depressive episode |  |  |  | 7.27 | (6.70, 7.90) | <0.0008* |
| F33 | Major depressive disorder, recurrent |  |  |  | 7.11 | (6.06, 8.34) | <0.0008* |
| F34.1 | Dysthymic disorder |  |  |  | 2.45 | (1.70, 3.52) | <0.0008* |
| **F40-F48** | **Anxiety, dissociative, stress-related, somatoform and other nonpsychotic mental disorders** | **5·69** | **(4·23, 7·64)** | **\*<0·0008** | **3·64** | **(3·43, 3·87)** | **<0·0008\*** |
| **F41·0** | **Panic disorder [episodic paroxysmal anxiety]** |  |  |  | **4·27** | **(3·35, 5·43)** | **<0·0008\*** |
| **F41·1** | **Generalized anxiety disorder** |  |  |  | **2·86** | **(2·54, 3·22)** | **<0·0008\*** |
| **F41·9** | **Anxiety disorder, unspecified** | **5·03** | **(3·36, 7·52)** | **\*<0·0008** | **3·47** | **(3·21, 3·74)** | **<0·0008\*** |
| **F42** | **Obsessive-compulsive disorder** |  |  |  | **3·27** | **(2·48, 4·32)** | **<0·0008\*** |
| **F43·1** | **Post-traumatic stress disorder (PTSD)** |  |  |  | **7·43** | **(6·16, 8·96)** | **<0·0008\*** |
| **F43·2** | **Adjustment disorders** | **7·04** | **(3·82, 12·97)** | **\*<0·0008** | **2·56** | **(2·28, 2·87)** | **<0·0008\*** |
| **F60-F69** | **Disorders of adult personality and behavior** | **3·67** | **(2·02, 6·68)** | **\*<0·0008** | **3·59** | **(3·11, 4·16)** | **<0·0008\*** |
| **F60·3** | **Borderline personality disorder** |  |  |  | **8·92** | **(5·73, 13·87)** | **<0·0008\*** |
| **F64** | **Gender identity disorders** |  |  |  | **5·82** | **(3·95, 8·58)** | **<0·0008\*** |
| **F70-F79** | **Intellectual Disabilities** |  |  |  | 1·28 | (0·99, 1·64) | 0·057 |
| **F84·0** | **Autistic disorder** | 2·68 | (1·50, 4·79) | 0·0009 | **2·08** | **(1·76, 2·44)** | **<0·0008\*** |
| **F84·5** | **Asperger's syndrome** |  |  |  | **3·01** | **(1·89, 4·78)** | **<0·0008\*** |
| **F90** | **Attention-deficit hyperactivity disorders** | **7·93** | **(5·75, 10·93)** | **\*<0·0008** | **2·58** | **(2·40, 2·78)** | **<0·0008\*** |
| **F91** | **Conduct disorders** | **11·50** | **(6·84, 19·34)** | **\*<0·0008** | **3·52** | **(3·14, 3·95)** | **<0·0008\*** |
| **MEDICAL DIAGNOSES** |  |  |  |  |  |  |  |
| **J45** | **Asthma** | **1·59** | **(1·28, 1·98)** | **\*<0·0008** | 1·01 | (0·94, 1·08) | 0·8581 |
| **E00-E07** | **Disorders of thyroid gland** |  |  |  | 1·00 | (0·82, 1·22) | 1 |
| **E08-E13** | **Diabetes mellitus** |  |  |  | 1·18 | (0·97, 1·43) | 0·0889 |
| **G40** | **Epilepsy and recurrent seizures** | 1·51 | (0·80, 2·85) | 0·20701 | 0·94 | (0·79, 1·12) | 0·4811 |
| **G47** | **Sleep disorders** | **2·22** | **(1·64, 3·00)** | **\*<0·0008** | **1·74** | **(1·59, 1·91)** | **<0·0008\*** |
| **G47·0** | **Insomnia** |  |  |  | **3·54** | **(2·97, 4·21)** | **<0·0008\*** |
| **G43** | **Migraine** | 1·00 | (0·45, 2·23) | 1 | 1·01 | (0·89, 1·15) | 0·8796 |

### Effects of COVID-19 on the Association with Suicidality and Self-Harm

#### Ages 0-12

During separate comparison of COVID-19(+) and COVID-19(-) children who all experienced SOSH, COVID-19 diagnosis itself was associated only with a pre-existing diagnosis of asthma (OR 1·75, 95% CI (1·41, 2·18), p<0·0008) (Supplementary Table 1). Among children diagnosed with SI/SA, COVID-19 diagnosis was associated with pre-existing diagnosis of asthma (OR 2·06, 95%CI (1·63, 2·61), p<0·0008) and insomnia (OR 3.01, 95%CI (1.70,5.34), p=0.0002 (Supplementary Table 2).

However, upon analysis of the variance of the odds ratios, the specific effect size of COVID-19 diagnosis was not significant for any pre-existing characteristic, for patients with SOSH or SI/SA.

#### Ages 13-21

In adolescents with SOSH, COVID-19 was associated with greater odds of the pre-existing psychiatric diagnoses ADHD (OR 1·35, 95%CI (1·23, 1·47), p<0·0008), conduct disorder (OR 1·34, 95%CI (1·19, 1·51), p<0·0008), adjustment disorders (OR 1·44, 95%CI (1·20, 1·74), p<0·0001), mood disorders (OR 1·26, 95%CI (1·12, 1·41), p<0·0001), anxiety disorders (OR 1·35, 95%CI (1·23, 1·48), p<0·0008), unspecified anxiety (OR 1·29, 95%CI (1·14, 1·47), p<0·0008), and depressive episode (OR 1·32, 95%CI (1·15, 1·52), p<0·0001) (Supplementary Table 3). In this group, there was higher odds of medical diagnoses including asthma (OR 1·36, 95%CI (1·24, 1·49), p<0·0008), sleep disorders (OR 1·35, 95%CI (1·19, 1·54), p<0·0008), insomnia (OR 1·56, 95%CI (1·21, 2·01), p<0·0006) and migraines (OR 1·81, 95%CI (1·44, 2·29), p<0·0008).

In adolescents with SI/SA, COVID-19 was associated with greater odds of the psychiatric risk factors ADHD (OR 1·25, 95%CI (1·18, 1·33), p<0·0008), conduct disorder (OR 1·28, 95%CI (1·18, 1·40), p<0·0008), bipolar disorder (OR 1·39, 95%CI (1·21, 1·59), p<0·0008), and unspecified anxiety (OR 1·14, 95% CI (1·08, 1·21), p<0·0008) (Supplementary Table 4).

Additionally, there were greater odds of socioeconomic factors (personal history of unspecified abuse in childhood (OR 2·57, 95%CI (1·85, 3·57), p<0·0008)) and medical diagnoses (asthma (OR 1·22, 95%CI (1·13, 1·31), p<0·0008) and diabetes mellitus (OR 1·66, 95%CI (1·34, 2·04), p<0·0008)).

In contrast, there were lower odds of socioeconomic risk factors related to education and literacy (OR 0·71, 95% CI (0·60, 0·83), p<0·0008), upbringing problems (OR 0·83, 95% CI (0·76, 0·90), p<0·0008), parent-child conflict (OR 0·65, 95% CI (0·56, 0·75), p<0·0008), history of childhood psychological abuse (OR 0·44, 95% CI (0·34, 0·56), p<0·0008); and psychiatric risk factors including mental disorders due to psychoactive substance use (OR 0·85, 95% CI (0·78, 0·93), p=0·0003), alcohol-related disorders (OR 0·65, 95% CI (0·53, 0·79), p<0·0008), gender identity disorder (OR 0·71, 95% CI (0·58, 0·86), p=0·0006), history of self-harm (OR 0·87, 95% CI (0·81, 0·94), p=0·0003), history of non-suicidal self-harm (OR 0·56, 95% CI (0·47, 0·68), p<0·0008), mood disorders (OR 0·83, 95% CI (0·80, 0·87), p<0·0008), generalized anxiety disorder (OR 0·86, 95% CI (0·79, 0·93), p=0·0005), depressive episode (OR 0·86, 95% CI (0·82, 0·90), p<0·0008), and recurrent major depressive disorder (OR 0·70, 95% CI (0·64, 0·75), p<0·0008).

In this age group, the effect size of COVID-19 was not significant for any pre-existing characteristic for patients with SOSH or SI/SA.

### Children vs. Adolescents

In patients with SOSH, patients diagnosed with COVID-19 had significantly greater odds of only one characteristic in children, and of several in adolescents (Table 14). In patients with SI/SA, patients with COVID-19 had significantly greater odds of two characteristics in children, and of several in adolescents (Table 15).

**Table 14.** Odds ratios (OR) of medical, psychiatric, and socioeconomic characteristics in patients with SOSH with or without COVID-19, with a comparison between OR for the 0-12 and 13-21 age groups. Bold denotes significantly greater odds. Italics denotes significantly lower odds. Statistical comparisons are represented by odds ratios (OR) with 95% confidence intervals (CI). Significant p-values are indicated with an asterisk. Characteristics observed in a total of 10 or fewer patients were excluded from comparison and are not included in this table.

| ICD-10 Code | Characteristic | 0-12 |  |  | 13-21 |  |  |
| --- | --- | --- | --- | --- | --- | --- | --- |
| SOCIOECONOMIC DIAGNOSES |  | OR | 95%CI | p value | OR | 95%CI | p value |
| Z55 | Problems related to education and literacy | 1.06 | (0.53, 2.11) | 0.8708 | 0.89 | (0.66, 1.20) | 0.4507 |
| Z59 | Problems related to housing and economic circumstances | 1.25 | (0.58, 2.69) | 0.5742 | 1.13 | (0.57, 2.21) | 0.7446 |
| Z60 | Problems related to social environment |  |  |  | 0.78 | (0.46, 1.32) | 0.3605 |
| Z62 | Problems related to upbringing | 0.90 | (0.65, 1.24) | 0.5252 | 1.15 | (0.91, 1.45) | 0.2564 |
| Z62.2 | Upbringing away from parents | 0.85 | (0.51, 1.41) | 0.5285 | 0.84 | (0.56, 1.27) | 0.4116 |
| Z62.81 | Personal history of abuse in childhood | 0.94 | (0.59, 1.52) | 0.8204 | 0.95 | (0.62, 1.47) | 0.8362 |
| Z62.810 | Personal history of physical and sexual abuse in childhood | 0.65 | (0.35, 1.20) | 0.1705 | 1.07 | (0.65, 1.76) | 0.8384 |
| Z62.82 | Parent-child conflict | 0.95 | (0.51, 1.76) | 0.8842 | 1.64 | (1.06, 2.53) | 0.0254 |
| Z63 | Other problems related to primary support group, including family circumstances | 1.23 | (0.87, 1.73) | 0.2546 | 1.37 | (1.06, 1.76) | 0.0141 |
| Z65 | Problems related to other psychosocial circumstances | 0.73 | (0.37, 1.47) | 0.3898 | 1.41 | (0.96, 2.08) | 0.081 |
| Z81 | Family history of mental and behavioral disorders | 1.07 | (0.74, 1.56) | 0.7175 | 1.02 | (0.78, 1.34) | 0.8978 |
| Z91.5 | Personal history of self-harm | 1.17 | (0.79, 1.73) | 0.4366 | 0.89 | (0.63, 1.25) | 0.502 |
| PSYCHIATRIC DIAGNOSES |  |  |  |  |  |  |  |
| F10-F19 | Mental and behavioral disorders due to psychoactive substance use |  |  |  | 0.91 | (0.62, 1.34) | 0.6345 |
| F12 | Cannabis related disorders |  |  |  | 0.71 | (0.40, 1.27) | 0.2528 |
| F17 | Nicotine dependence |  |  |  | 1.33 | (0.68, 2.61) | 0.4067 |
| F20-F29 | Schizophrenia, schizotypal, delusional, and other non-mood psychotic disorders |  |  |  | 1.36 | (0.92, 2.00) | 0.121 |
| F29 | Unspecified psychosis not due to a substance or known physiological condition |  |  |  | 1.62 | (1.06, 2.49) | 0.027 |
| **F30-F39** | **Mood [affective] disorders** | 0.81 | (0.65, 1.01) | 0.0565 | **1.26** | **(1.12, 1.41)** | **0.0001\*** |
| **F31** | **Bipolar disorder** |  |  |  | 1.34 | (0.95, 1.90) | 0.0971 |
| **F32** | **Depressive episode** | 0.68 | (0.51, 0.91) | 0.0088 | 1.32 | (1.15, 1.52) | *0.0001 |
| **F33** | **Major depressive disorder, recurrent** | 0.95 | (0.50, 1.79) | 0.8812 | 1.00 | (0.75, 1.34) | 1 |
| **F34.1** | **Dysthymic disorder** |  |  |  | 0.76 | (0.44, 1.32) | 0.3332 |
| **F40-F48** | **Anxiety, dissociative, stress-related, somatoform and other nonpsychotic mental disorders** | 1.09 | (0.90, 1.32) | 0.3682 | **1.35** | **(1.23, 1.48)** | **<0.0008\*** |
| **F41.0** | **Panic disorder [episodic paroxysmal anxiety]** |  |  |  | 1.40 | (0.88, 2.24) | 0.1594 |
| **F41.1** | **Generalized anxiety disorder** | 0.92 | (0.59, 1.45) | 0.745 | 1.18 | (0.97, 1.44) | 0.0964 |
| **F41.9** | **Anxiety disorder, unspecified** | 1.08 | (0.84, 1.38) | 0.5763 | **1.29** | **(1.14, 1.47)** | **<0.0008\*** |
| **F42** | **Obsessive-compulsive disorder** |  |  |  | 1.31 | (0.89, 1.94) | 0.1712 |
| **F43.1** | **Post-traumatic stress disorder (PTSD)** | 1.09 | (0.72, 1.66) | 0.6846 | 1.19 | (0.92, 1.54) | 0.1892 |
| **F43.2** | **Adjustment disorders** | 1.12 | (0.80, 1.56) | 0.5093 | **1.44** | **(1.20, 1.74)** | **0.0001\*** |
| **F60-F69** | **Disorders of adult personality and behavior** | 0.83 | (0.57, 1.21) | 0.3447 | 1.20 | (1.02, 1.41) | 0.0241 |
| **F60.3** | **Borderline personality disorder** |  |  |  | 1.11 | (0.77, 1.61) | 0.5821 |
| **F64** | **Gender identity disorders** |  |  |  | 1.18 | (0.53, 2.64) | 0.6965 |
| **F70-F79** | **Intellectual Disabilities** |  |  |  | 1.07 | (0.70, 1.63) | 0.7603 |
| **F84.0** | **Autistic disorder** | 0.66 | (0.45, 0.96) | 0.0305 | 1.21 | (0.97, 1.50) | 0.0897 |
| **F84.5** | **Asperger's syndrome** |  |  |  | 1.18 | (0.74, 1.86) | 0.4953 |
| **F90** | **Attention-deficit hyperactivity disorders** | 1.15 | (0.95, 1.38) | 0.1543 | **1.35** | **(1.23, 1.47)** | **<0.0008\*** |
| **F91** | **Conduct disorders** | 1.14 | (0.90, 1.45) | 0.2812 | **1.34** | **(1.19, 1.51)** | **<0.0008\*** |
| **MEDICAL DIAGNOSES** |  |  |  |  |  |  |  |
| **J45** | **Asthma** | **1.75** | **(1.41, 2.18)** | **\*<0.0008** | **1.36** | **(1.24, 1.49)** | **<0.0008\*** |
| **E00-E07** | **Disorders of thyroid gland** |  |  |  | 1.13 | (0.80, 1.59) | 0.4954 |
| **E08-E13** | **Diabetes mellitus** |  |  |  | 1.52 | (1.09, 2.12) | 0.0134 |
| **G40** | **Epilepsy and recurrent seizures** | 1.54 | (0.86, 2.76) | 0.1487 | 1.24 | (0.96, 1.61) | 0.102 |
| **G47** | **Sleep disorders** | 1.36 | (1.05, 1.76) | 0.0215 | **1.35** | **(1.19, 1.54)** | **<0.0008\*** |
| **G47.0** | **Insomnia** | 1.43 | (0.93, 2.21) | 0.1036 | **1.56** | **(1.21, 2.01)** | **0.0006\*** |
| **G43** | **Migraine** | 0.93 | (0.43, 1.98) | 0.8571 | **1.81** | **(1.44, 2.29)** | **<0.0008\*** |

**Table 15.**
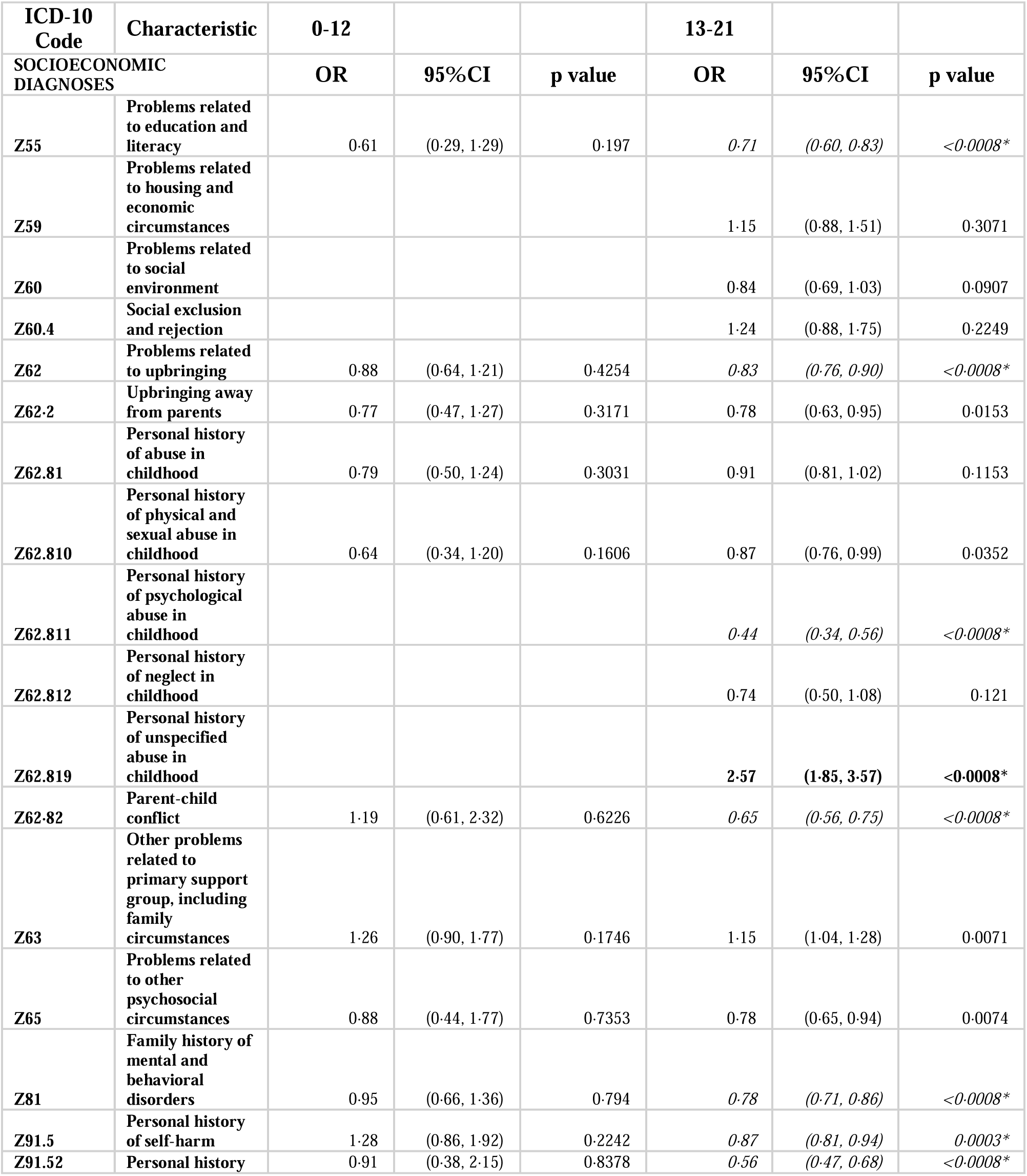
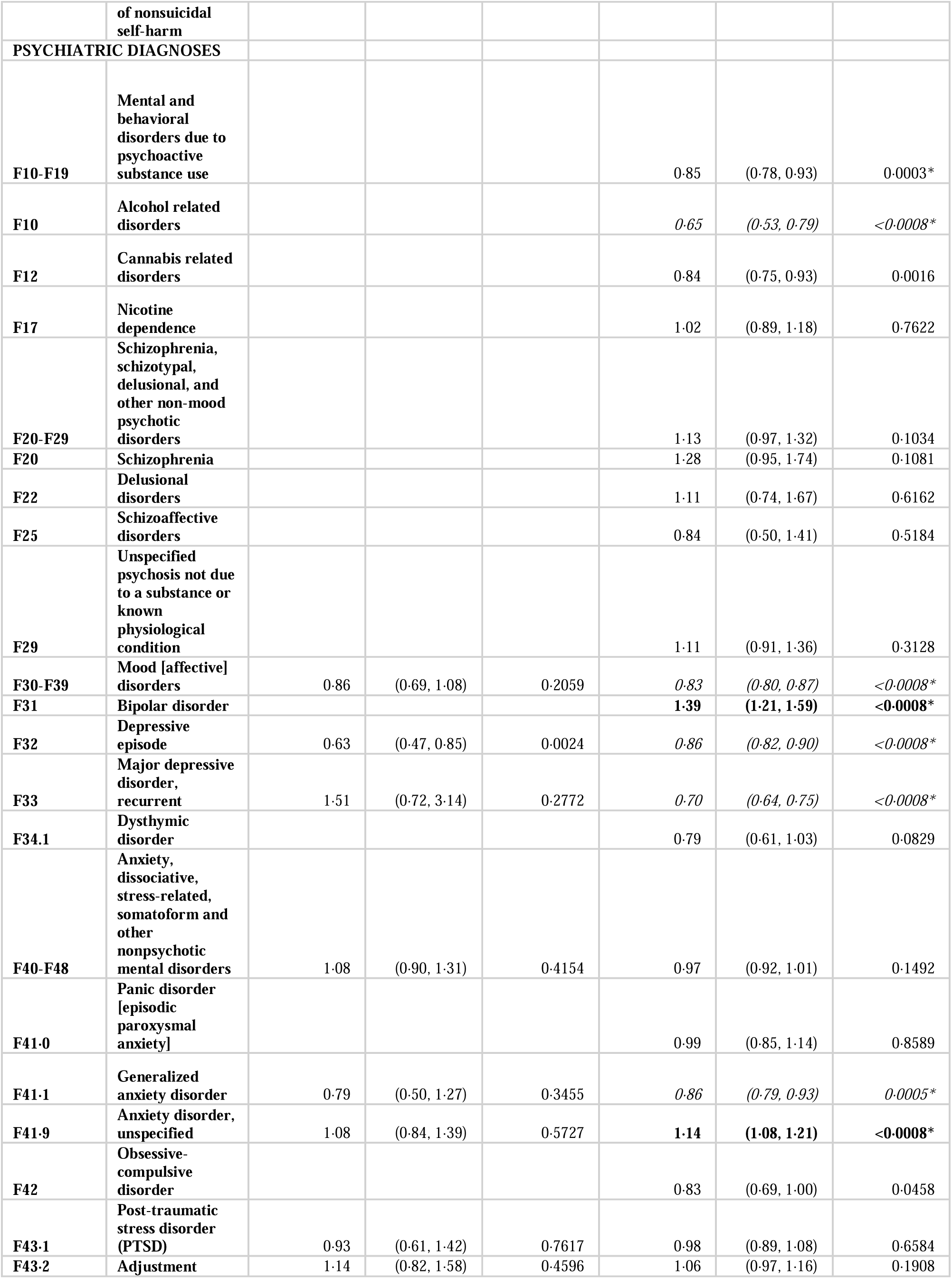
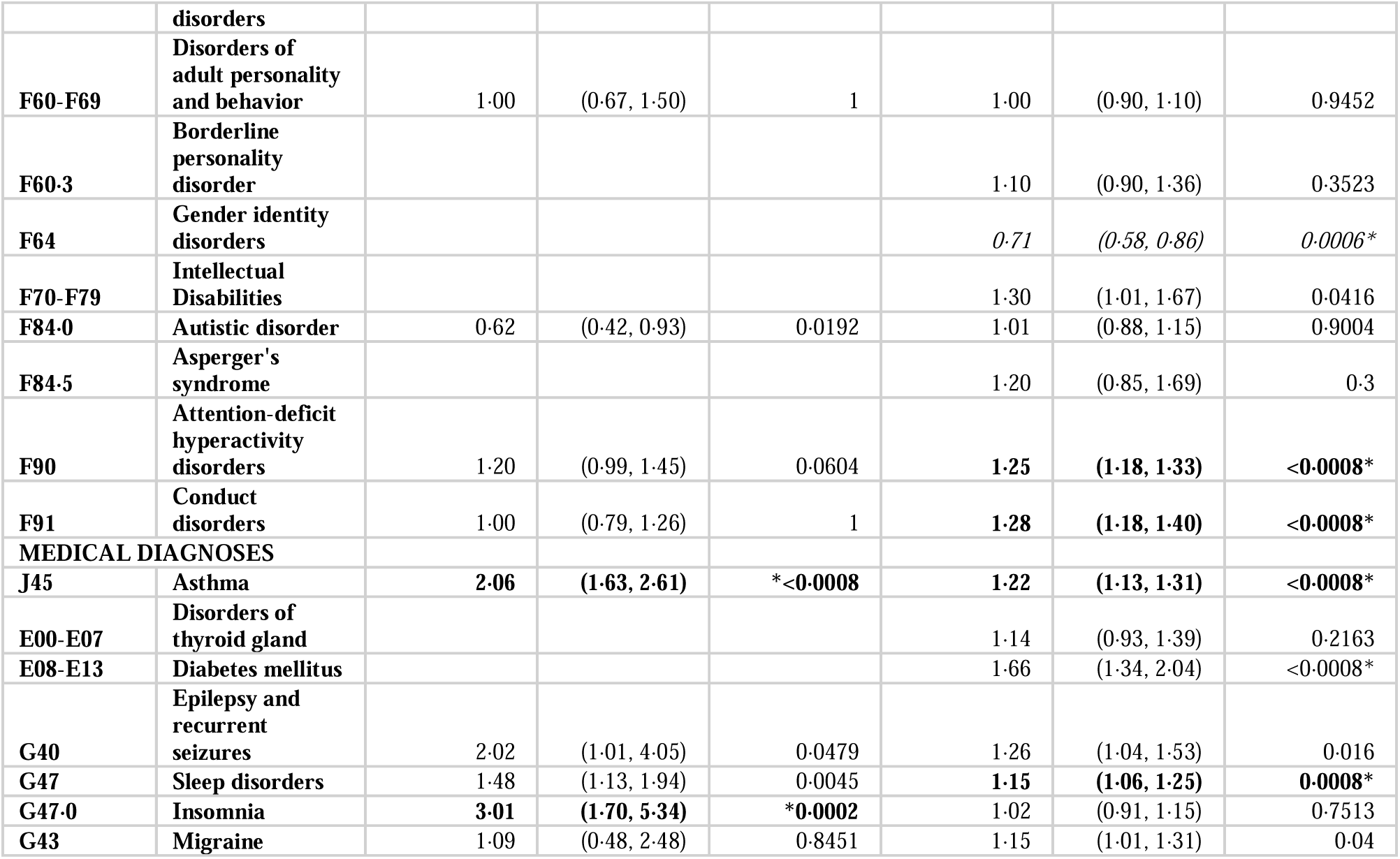
Odds ratios (OR) of medical, psychiatric, and socioeconomic characteristics in patients with SI/SA with or without COVID-19, with a comparison between OR for the 0-12 and 13-21 age groups. Bold denotes significantly greater odds. Italics denotes significantly lower odds. Statistical comparisons are represented by odds ratios (OR) with 95% confidence intervals (CI). Significant p-values are indicated with an asterisk. Characteristics observed in a total of 10 or fewer patients were excluded from comparison and are not included in this table.

## DISCUSSION

This is the first study to specifically examine this number of risk factors for suicidality and self-harm in both children and adolescents in relation to COVID-19 diagnosis and also examine the effect of COVID-19. Though we examined known risk factors, we observed a marked variety of ORs across these pre-existing characteristics: for some, we observed significant association with SOSH and SI/A, and for others did not observe such strong association.

In both children and adolescents, patients with SOSH and SI/SA had a greater odds of a prior COVID-19 diagnosis than patients without. In these two age groups, SOSH and SI/SA were associated with several pre-existing psychiatric disorders, medical diagnoses, and socioeconomic factors previously described in the literature. Problems with primary support group, adult personality disorders, GAD, thyroid disorders, and insomnia were significant only in COVID-19+ children with SOSH; while upbringing problems, anxiety and other nonpsychotic disorders, sleep disorders, and autistic disorder were significant only in COVID-19-children with SOSH. In children with SI/SA, problems with primary support group, adult personality disorders, and asthma were significant only in COVID-19+ patients; while autistic disorder was significant only in COVID-19-patients. In adolescents with SOSH, problems related to education and literacy were significant only in COVID-19-patients, while parent-child conflict was only significant in COVID-19+ patients. In adolescents with SI/SA, asthma was only significant in COVID-19-patients.

This study reaffirms the associations between factors related to suicidality that exist independently of the pandemic context. These include psychiatric diagnoses like ADHD and conduct disorder^(5)^, medical diagnoses like insomnia or sleep disorders^(23)^, and socioeconomic factors like childhood abuse^(24)^. In contrast to prior literature, this study does not consistently demonstrate an association between SOSH and medical risk factors like diabetes and asthma.

We found a greater odds of COVID-19 in patients with suicidality than without but found little difference between the factors associated with suicidality in COVID-19(+) and COVID-19(-) patients.

Greater suicidality rates have been observed in COVID-19-infected patients both before and after SARS-CoV-2 diagnosis^(8, 9)^. Development of SOSH after COVID-19 diagnosis could be associated with COVID-19-related inflammation^(12)^, potentially related to the emerging relationship between inflammation and depression^(25)^ and between inflammation and suicide^(26)^. This study has several strengths. Replicating associations between SOSH and previously studied risk factors in a large sample documents generalizability. Separating child and adolescent populations addresses variability between age groups, including the possibility of unique risk factors. Additionally, our focus on suicidality after COVID-19 diagnosis attempts to examine associations of the disease itself, rather than the psychological impact of the pandemic.

Regarding limitations, this study used TriNetX, which is sourced from electronic medical records and examines diagnoses based on ICD codes. The accuracy of EMR data may vary based on documentation practices. Providers may be less likely to document socioeconomic factors like abuse or homelessness and may not attach corresponding ICD codes. The database likely underestimates the true set of patients with these socioeconomic concerns.

This study controlled for a limited set of covariates through propensity score-matching. Due to platform limitations (inability to balance for greater than a set number of covariates for very large samples), we restricted propensity-score matching to demographic factors (age, sex, race, ethnicity), and so the present study fails to account for the confounding effect of pre-existing psychiatric disorders (e.g. depression), medical history, and socioeconomic factors. Additional confounders may remain. As we controlled for demographic factors, our study also does not address disparities in SOSH or risk factors across racial, ethnic, and sex groups.

While this study examined SOSH (SI/SA/SIB) and suicidality (SI/SA) separately, it does not individually examine SI, SA, and SIB, which may have different risk factor profiles. Furthermore, this study does not account for variability in the expression of COVID-19 that may impact suicide risk, including severity of COVID-19 infection, need for hospitalization, burden of COVID-19 symptoms, and persistence of symptoms after infection^(27)^.

Our study is the first to examine risk factors for suicidality and self-harm associated with COVID-19 diagnosis and infection in a large sample of children and adolescents; demonstrates increased odds of preceding COVID-19 diagnosis in patients with suicidality and self-harm; and demonstrates minimal association between COVID-19 and pre-existing risk factors in children, but association with several psychiatric, medical, and socioeconomic factors in adolescents.

## Author Contribution Statement

All listed authors have made substantial contributions to this work through all of most of the following: initial design, data acquisition and analysis, drafting, and review. All authors have reviewed this work and approve of the final version submitted for publication. All authors agree to take accountability for all aspects of this work and ensure that any questions related to the accuracy or integrity of any part of the work are appropriately investigated and/or resolved.

## AI Statement

No AI tools were used in the writing or editing of this manuscript.

## Data Sharing Statement

The individual patient-level electronic health record data in the TriNetX database cannot be shared, as they are the property of TriNetX and may only be accessed through direct contract with the company. Individual-level data was not accessed by the researchers involved in this study. Summary data and results are provided here in the manuscript’s Tables and Supplementary Tables.

