## supplementary file for "Risk Factors for Suicidal Behavior in Youth and the Impact of SARS-CoV-2 Infection: A Retrospective Case-Control Study"

**Supplemental Information**

**COVID-19 DIAGNOSIS**

We defined COVID-19 diagnosis based on either positive laboratory tests or COVID-19 diagnostic codes.

Laboratory tests included the following: 9088, SARS coronavirus 2 and related RNA [Presence]; 944661-6, SARS-CoV-2 (COVID-19) Ab [Interpretation] in Serum or Plasma; 94533-7, SARS-CoV-2 (COVID-19) N gene [Presence] in Respiratory specimen by NAA with probe detection; 87811, Infectious agent antigen detection by immunoassay with direct optical (ie visual) observation; severe acute respiratory syndrome coronavirus 2 (SARSCoV-2) (Coronavirus disease [COVID-19]); 94500-6, SARS-CoV-2 (COVID-19) RNA [Presence] in Respiratory specimen by NAA with probe detection; 94563-4, SARS-CoV (COVID-19) IgG Ab [Presence] in Serum or Plasma by Immunoassay; 9089, SARS coronavirus 2 IgG IgM Ab [Presence] in Serum or Plasma; 94534-5, SARS-CoV-2 (COVID-19) RdRp gene [Presence] in Respiratory specimen by NAA with probe detection; 94316-7, SARS-CoV-2 (COVID-19) N gene [Presence] in Unspecified specimen by NAA with probe detection; 94558-4, SARS-CoV-2 (COVID-19) Ag [Presence] in Respiratory specimen by rapid immunoassay; 94507-1, SARS-CoV-2 (COVID-19) IgG Ab [Presence] in Serum, Plasma or Blood by Rapid immunoassay; 94759-8, SARS-CoV-2 (COVID-19) RNA [Presence] in Nasopharynx by NAA with probe detection; 95209-3, SARS-CoV-+SARS-CoV-2 (COVID-19) Ag [Presence] in Respiratory specimen by Rapid immunoassay; 94565-9, SARS-CoV-2 (COVID-19) RNA [Presence] in Nasopharynx by NAA with non-probe detection; 94505-5, SARS-CoV-2 (COVID-19) IgG Ab [units/volume] in Serum or Plasma by immunoassay; 94762-2, SARS-CoV-2 (COVID-19) Ab [Presence] in Serum or Plasma by Immunoassay; 95608-6, SARS-CoV-2 (COVID-19) RNA [Presence] in Respiratory specimen by NAA with probe detection; 9455-2, SARS-CoV-2 (COVID-19) ORF1ab region [Presence] in Respiratory specimen by NAA with probe detection; 94564-2, SARS-CoV-2 (COVID-19) IgM Ab [Presence] in Serum or Plasma by Immunoassay; 94314-2, SARS-CoV-2 (COVID-19) RdRp gene [Presence] in Unspecified specimen by NAA with probe detection; 94547-7, SARS-CoV-2 (COVID-19) IgG+IgM Ab [Presence] in Serum or Plasma by Immunoassay; 95406-5, SARS-CoV-2 (COVID-19) RNA [Presence] in Nose by NAA with probe detection; 94769-7, SARS-CoV-2 (COVID-19) Ab [units/volume] in Serum or Plasma by Immunoassay; 94531-1, SARS-CoV-2 (COVID-19) RNA panel – Respiratory specimen by NAA with probe detection; 94508-9, SARS-CoV-2 (COVID-19) IgM Ab [Presence] in Serum, Plasma or Blood by Rapid immunoassay; 94760-6, SARS-CoV-2 (COVID-19) N gene [Presence] in Nasopharynx by NAA with probe detection; 94562-6, SARS-CoV-2 (COVID-19) IgA Ab [Presence] in Serum or Plasma by Immunoassay; 94506-3, SARS-CoV-2 (COVID-19) IgM Ab [units/volume] in Serum or Plasma by Immunoassay; 87635, Infectious agent detection by nucleic acid (DNA or RNA); severe acute respiratory syndrome coronavirus 2 (SARS-CoV-2) (Coronavirus disease [COVID-19]), amplified probe technique; 94306-8, SARS-CoV-2 (COVID-19) RNA panel – unspecified specimen by NAA with probe detection; 86413, Severe acute respiratory syndrome coronavirus 2 (SARS-CoV-2) (Coronavirus disease [COVID-19]), antibody, quantitative; 94503-0, SARS-CoV-2 (COVID-19) Ab panel – Serum, Plasma or Blood by Rapid immunoassay.

COVID-19 diagnosis included: J12.82, Pneumonia due to COVID-19; U07.2, COVID-19, virus not identified (WHO); U07.1, COVID-19.

**VARIANCE OF ODDS RATIO CALCULATION**

We determined the variance of the odds ratios four times: for the 0-12 SOSH analysis, 0-12 SI/SA analysis, 13-21 SOSH analysis, and 13-21 SI/SA analysis. Formulas were drawn from the Cochrane Handbook for Systematic Reviews of Interventions (Higgins and Thomas, 2023), and from (Altman and Bland, 2011).

For the 0-12 SOSH analysis:

For each of the 53 risk factors, we determined the odds ratio obtained from the COVID+ cohort (COVID-19 with SOSH v. COVID-19 without SOSH), the confidence interval lower-limit, and confidence interval upper-limit; and the odds ratio from the COVID- cohort (No COVID-19 with SOSH v. No COVID-29 without SOSH), confidence interval lower-limit, and confidence interval upper-limit. Here, “OR1” is “OR for the COVID+ cohort,” and “OR2” is “OR for the COVID- cohort.”

1. We calculated the ratio of the ratios:

OR Ratio= OR1/OR2

Or

OR Ratio = OR for COVID+ cohort / OR for COVID- cohort

2. We separately calculated the natural log of OR1 and OR2: ln(OR1) and ln(OR2).

3. We calculated the individual variance of ln(OR1) and ln(OR2), according to the formula:

SE = (upper limit – lower limit)/(2x1.96)

Variance =  SE^2

Variance =((ln(upper limit of CI) – ln(lower limit of CI)) / (2x1.96))^2.

4. We calculated the variance of the log of the OR ratio with the formula:

Variance of log(OR Ratio) = Variance of log(OR1) + Variance of log(OR2)

5. We calculated the 95% CI for the log(OR ratio) with the formula:

CI upper limit = exp(ln(OR Ratio) + 1.96 x sqrt(variance of log(OR ratio)))

CI lower limit = exp(ln(OR ratio) – 1.96 x sqrt(variance of log(OR ratio)))

6. We calculated the Z score with the formula:

Z-score = (ln(OR Ratio)) / (sqrt(variance of log(OR ratio)))

7. We calculated p value with the formula, according to (Altman and Bland, 2011):

P = exp((-0.717*abs(Z score))-(0.416*(abs(Z score))^2))

**Supplementary Tables**

| **ICD-10 Code** | **Characteristic** | **SOSH(+), COVID-19(+)** | | **SOSH(+), COVID-19(-)** | | **OR** | **95%CI** | **P value** |
| --- | --- | --- | --- | --- | --- | --- | --- | --- |
|  |  | **N** | **%** | **N** | **%** |  |  |  |
| **SOCIOECONOMIC DIAGNOSES** | |  | |  | |  |  |  |
| **Z55** | **Problems related to education and literacy** | 17 | 1·25% | 16 | 1·18% | 1·06 | (0·53, 2·11) | 0·8708 |
| **Z59** | **Problems related to housing and economic circumstances** | 15 | 1·11% | 12 | 0·88% | 1·25 | (0·58, 2·69) | 0·5742 |
| **Z62** | **Problems related to upbringing** | 76 | 5·60% | 84 | 6·19% | 0·90 | (0·65, 1·24) | 0·5252 |
| **Z62·2** | **Upbringing away from parents** | 28 | 2·06% | 33 | 2·43% | 0·85 | (0·51, 1·41) | 0·5285 |
| **Z62.81** | **Personal history of abuse in childhood** | 34 | 2·51% | 36 | 2·65% | 0·94 | (0·59, 1·52) | 0·8204 |
| **Z62.810** | **Personal history of physical and sexual abuse in childhood** | 17 | 1·25% | 26 | 1·92% | 0·65 | (0·35, 1·20) | 0·1705 |
| **Z62·82** | **Parent-child conflict** | 20 | 1·47% | 21 | 1·55% | 0·95 | (0·51, 1·76) | 0·8842 |
| **Z63** | **Other problems related to primary support group, including family circumstances** | 74 | 5·45% | 61 | 4·50% | 1·23 | (0·87, 1·73) | 0·2546 |
| **Z65** | **Problems related to other psychosocial circumstances** | 14 | 1·03% | 19 | 1·40% | 0·73 | (0·37, 1·47) | 0·3898 |
| **Z81** | **Family history of mental and behavioral disorders** | 60 | 4·42% | 56 | 4·13% | 1·07 | (0·74, 1·56) | 0·7175 |
| **Z91.5** | **Personal history of self-harm** | 57 | 4·20% | 49 | 3·61% | 1·17 | (0·79, 1·73) | 0·4366 |
| **PSYCHIATRIC DIAGNOSES** | |  |  |  |  |  |  |  |
| **F30-F39** | **Mood [affective] disorders** | 167 | 12·31% | 201 | 14·81% | 0·81 | (0·65, 1·01) | 0·0565 |
| **F32** | **Depressive episode** | 83 | 6·12% | 119 | 8·77% | 0·68 | (0·51, 0·91) | 0·0088 |
| **F33** | **Major depressive disorder, recurrent** | 19 | 1·40% | 20 | 1·47% | 0·95 | (0·50, 1·79) | 0·8812 |
| **F40-F48** | **Anxiety, dissociative, stress-related, somatoform and other nonpsychotic mental disorders** | 281 | 20·71% | 262 | 19·31% | 1·09 | (0·90, 1·32) | 0·3682 |
| **F41·1** | **Generalized anxiety disorder** | 38 | 2·80% | 41 | 3·02% | 0·92 | (0·59, 1·45) | 0·745 |
| **F41·9** | **Anxiety disorder, unspecified** | 140 | 10·32% | 131 | 9·65% | 1·08 | (0·84, 1·38) | 0·5763 |
| **F43·1** | **Post-traumatic stress disorder (PTSD)** | 48 | 3·54% | 44 | 3·24% | 1·09 | (0·72, 1·66) | 0·6846 |
| **F43·2** | **Adjustment disorders** | 78 | 5·75% | 70 | 5·16% | 1·12 | (0·80, 1·56) | 0·5093 |
| **F60-F69** | **Disorders of adult personality and behavior** | 52 | 3·83% | 62 | 4·57% | 0·83 | (0·57, 1·21) | 0·3447 |
| **F84·0** | **Autistic disorder** | 47 | 3·46% | 70 | 5·16% | 0·66 | (0·45, 0·96) | 0·0305 |
| **F90** | **Attention-deficit hyperactivity disorders** | 293 | 21·59% | 263 | 19·38% | 1·15 | (0·95, 1·38) | 0·1543 |
| **F91** | **Conduct disorders** | 164 | 12·09% | 146 | 10·76% | 1·14 | (0·90, 1·45) | 0·2812 |
| **MEDICAL DIAGNOSES** | |  |  |  |  |  |  |  |
| **J45** | **Asthma** | 244 | 17·98% | 151 | 11·13% | 1·75 | (1·41, 2·18) | *<0·0008 |
| **G40** | **Epilepsy and recurrent seizures** | 29 | 2·14% | 19 | 1·40% | 1·54 | (0·86, 2·76) | 0·1487 |
| **G47** | **Sleep disorders** | 145 | 10·69% | 110 | 8·11% | 1·36 | (1·05, 1·76) | 0·0215 |
| **G47·0** | **Insomnia** | 51 | 3·76% | 36 | 2·65% | 1·43 | (0·93, 2·21) | 0·1036 |
| **G43** | **Migraine** | 13 | 0·96% | 14 | 1·03% | 0·93 | (0·43, 1·98) | 0·8571 |

**Supplementary Table 1.** Prevalence and odds ratios of medical/psychiatric/socioeconomic characteristics in patients 0-12 with or without COVID-19 among SOSH+ patients**.** Statistical comparisons are represented by odds ratios (OR) with 95% confidence intervals (CI)**.** Significant p-values are indicated with an asterisk (*)**.**

| **ICD-10 Code** | **Characteristic** | **SI/SA(+), COVID-19(+)** | | **SI/SA(+), COVID-19(-)** | | **OR** | **95%CI** | **P value** |
| --- | --- | --- | --- | --- | --- | --- | --- | --- |
|  |  | **N** | **%** | **N** | **%** |  |  |  |
| **SOCIOECONOMIC DIAGNOSES** | |  |  |  |  |  |  |  |
| **Z55** | **Problems related to education and literacy** | 11 | 0·84% | 18 | 1·38% | 0·61 | (0·29, 1·29) | 0·197 |
| **Z62** | **Problems related to upbringing** | 76 | 5·83% | 86 | 6·60% | 0·88 | (0·64, 1·21) | 0·4254 |
| **Z62·2** | **Upbringing away from parents** | 28 | 2·15% | 36 | 2·76% | 0·77 | (0·47, 1·27) | 0·3171 |
| **Z62.81** | **Personal history of abuse in childhood** | 34 | 2·61% | 43 | 3·30% | 0·79 | (0·50, 1·24) | 0·3031 |
| **Z62.810** | **Personal history of physical and sexual abuse in childhood** | 16 | 1·23% | 25 | 1·92% | 0·64 | (0·34, 1·20) | 0·1606 |
| **Z62·82** | **Parent-child conflict** | 19 | 1·46% | 16 | 1·23% | 1·19 | (0·61, 2·32) | 0·6226 |
| **Z63** | **Other problems related to primary support group, including family circumstances** | 81 | 6·22% | 65 | 4·99% | 1·26 | (0·90, 1·77) | 0·1746 |
| **Z65** | **Problems related to other psychosocial circumstances** | 15 | 1·15% | 17 | 1·30% | 0·88 | (0·44, 1·77) | 0·7353 |
| **Z81** | **Family history of mental and behavioral disorders** | 60 | 4·60% | 63 | 4·83% | 0·95 | (0·66, 1·36) | 0·794 |
| **Z91.5** | **Personal history of self-harm** | 56 | 4·30% | 44 | 3·38% | 1·28 | (0·86, 1·92) | 0·2242 |
| **Z91.52** | **Personal history of nonsuicidal self-harm** | 10 | 0·77% | 11 | 0·84% | 0·91 | (0·38, 2·15) | 0·8378 |
| **PSYCHIATRIC DIAGNOSES** | |  |  |  |  |  |  |  |
| **F30-F39** | **Mood [affective] disorders** | 162 | 12·43% | 184 | 14·12% | 0·86 | (0·69, 1·08) | 0·2059 |
| **F32** | **Depressive episode** | 76 | 5·83% | 117 | 8·98% | 0·63 | (0·47, 0·85) | 0·0024 |
| **F33** | **Major depressive disorder, recurrent** | 18 | 1·38% | 12 | 0·92% | 1·51 | (0·72, 3·14) | 0·2772 |
| **F40-F48** | **Anxiety, dissociative, stress-related, somatoform and other nonpsychotic mental disorders** | 273 | 20·95% | 256 | 19·65% | 1·08 | (0·90, 1·31) | 0·4154 |
| **F41·1** | **Generalized anxiety disorder** | 32 | 2·46% | 40 | 3·07% | 0·79 | (0·50, 1·27) | 0·3455 |
| **F41·9** | **Anxiety disorder, unspecified** | 138 | 10·59% | 129 | 9·90% | 1·08 | (0·84, 1·39) | 0·5727 |
| **F43·1** | **Post-traumatic stress disorder (PTSD)** | 44 | 3·38% | 47 | 3·61% | 0·93 | (0·61, 1·42) | 0·7617 |
| **F43·2** | **Adjustment disorders** | 80 | 6·14% | 71 | 5·45% | 1·14 | (0·82, 1·58) | 0·4596 |
| **F60-F69** | **Disorders of adult personality and behavior** | 49 | 3·76% | 49 | 3·76% | 1·00 | (0·67, 1·50) | 1 |
| **F84·0** | **Autistic disorder** | 42 | 3·22% | 66 | 5·07% | 0·62 | (0·42, 0·93) | 0·0192 |
| **F90** | **Attention-deficit hyperactivity disorders** | 293 | 22·49% | 254 | 19·49% | 1·20 | (0·99, 1·45) | 0·0604 |
| **F91** | **Conduct disorders** | 163 | 12·51% | 163 | 12·51% | 1·00 | (0·79, 1·26) | 1 |
| **MEDICAL DIAGNOSES** | |  |  |  |  |  |  |  |
| **J45** | **Asthma** | 229 | 17·57% | 122 | 9·36% | 2·06 | (1·63, 2·61) | *<0·0008 |
| **G40** | **Epilepsy and recurrent seizures** | 24 | 1·84% | 12 | 0·92% | 2·02 | (1·01, 4·05) | 0·0479 |
| **G47** | **Sleep disorders** | 140 | 10·74% | 98 | 7·52% | 1·48 | (1·13, 1·94) | 0·0045 |
| **G47·0** | **Insomnia** | 47 | 3·61% | 16 | 1·23% | 3·01 | (1·70, 5·34) | *0·0002 |
| **G43** | **Migraine** | 12 | 0·92% | 11 | 0·84% | 1·09 | (0·48, 2·48) | 0·8451 |

**Supplementary Table 2.** Prevalence and odds ratios of medical/psychiatric/socioeconomic characteristics in patients 0-12 with or without COVID-19 among SI/SA+ patients**.** Statistical comparisons are represented by odds ratios (OR) with 95% confidence intervals (CI)**.** Significant p-values are indicated with an asterisk (*)**.**

| **ICD-10 Code** | **Characteristic** | **SOSH(+), COVVID-19(+)** | | **SOSH(+), COVID-19(-)** | | **OR** | **95%CI** | **P value** |
| --- | --- | --- | --- | --- | --- | --- | --- | --- |
|  |  | **N** | **%** | **N** | **%** |  |  |  |
| **SOCIOECONOMIC DIAGNOSES** | |  |  |  |  |  |  |  |
| **Z55** | **Problems related to education and literacy** | 80 | 0·52% | 90 | 0·59% | 0·89 | (0·66, 1·20) | 0·4507 |
| **Z59** | **Problems related to housing and economic circumstances** | 18 | 0·12% | 16 | 0·10% | 1·13 | (0·57, 2·21) | 0·7446 |
| **Z60** | **Problems related to social environment** | 25 | 0·16% | 32 | 0·21% | 0·78 | (0·46, 1·32) | 0·3605 |
| **Z60.4** | **Social exclusion and rejection** |  |  |  |  |  |  |  |
| **Z62** | **Problems related to upbringing** | 149 | 0·97% | 130 | 0·85% | 1·15 | (0·91, 1·45) | 0·2564 |
| **Z62·2** | **Upbringing away from parents** | 42 | 0·27% | 50 | 0·33% | 0·84 | (0·56, 1·27) | 0·4116 |
| **Z62.81** | **Personal history of abuse in childhood** | 40 | 0·26% | 42 | 0·27% | 0·95 | (0·62, 1·47) | 0·8362 |
| **Z62.810** | **Personal history of physical and sexual abuse in childhood** | 32 | 0·21% | 30 | 0·20% | 1·07 | (0·65, 1·76) | 0·8384 |
| **Z62·82** | **Parent-child conflict** | 54 | 0·35% | 33 | 0·22% | 1·64 | (1·06, 2·53) | 0·0254 |
| **Z63** | **Other problems related to primary support group, including family circumstances** | 146 | 0·95% | 107 | 0·70% | 1·37 | (1·06, 1·76) | 0·0141 |
| **Z65** | **Problems related to other psychosocial circumstances** | 62 | 0·41% | 44 | 0·29% | 1·41 | (0·96, 2·08) | 0·081 |
| **Z81** | **Family history of mental and behavioral disorders** | 105 | 0·69% | 103 | 0·67% | 1·02 | (0·78, 1·34) | 0·8978 |
| **Z91.5** | **Personal history of self-harm** | 64 | 0·42% | 72 | 0·47% | 0·89 | (0·63, 1·25) | 0·502 |
| **PSYCHIATRIC DIAGNOSES** | |  |  |  |  |  |  |  |
| **F10-F19** | **Mental and behavioral disorders due to psychoactive substance use** | 49 | 0·32% | 54 | 0·35% | 0·91 | (0·62, 1·34) | 0·6345 |
| **F12** | **Cannabis related disorders** | 20 | 0·13% | 28 | 0·18% | 0·71 | (0·40, 1·27) | 0·2528 |
| **F17** | **Nicotine dependence** | 20 | 0·13% | 15 | 0·10% | 1·33 | (0·68, 2·61) | 0·4067 |
| **F20-F29** | **Schizophrenia, schizotypal, delusional, and other non-mood psychotic disorders** | 61 | 0·40% | 45 | 0·29% | 1·36 | (0·92, 2·00) | 0·121 |
| **F29** | **Unspecified psychosis not due to a substance or known physiological condition** | 55 | 0·36% | 34 | 0·22% | 1·62 | (1·06, 2·49) | 0·027 |
| **F30-F39** | **Mood [affective] disorders** | 668 | 4·37% | 535 | 3·50% | 1·26 | (1·12, 1·41) | ·0001* |
| **F31** | **Bipolar disorder** | 75 | 0·49% | 56 | 0·37% | 1·34 | (0·95, 1·90) | 0·0971 |
| **F32** | **Depressive episode** | 460 | 3·01% | 350 | 2·29% | 1·32 | (1·15, 1·52) | *0·0001 |
| **F33** | **Major depressive disorder, recurrent** | 91 | 0·60% | 91 | 0·60% | 1·00 | (0·75, 1·34) | 1 |
| **F34.1** | **Dysthymic disorder** | 22 | 0·14% | 29 | 0·19% | 0·76 | (0·44, 1·32) | 0·3332 |
| **F40-F48** | **Anxiety, dissociative, stress-related, somatoform and other nonpsychotic mental disorders** | 1066 | 6·97% | 806 | 5·27% | 1·35 | (1·23, 1·48) | <0·0008* |
| **F41·0** | **Panic disorder [episodic paroxysmal anxiety]** | 42 | 0·27% | 30 | 0·20% | 1·40 | (0·88, 2·24) | 0·1594 |
| **F41·1** | **Generalized anxiety disorder** | 216 | 1·41% | 183 | 1·20% | 1·18 | (0·97, 1·44) | 0·0964 |
| **F41·9** | **Anxiety disorder, unspecified** | 577 | 3·77% | 450 | 2·94% | 1·29 | (1·14, 1·47) | <0·0008* |
| **F42** | **Obsessive-compulsive disorder** | 59 | 0·39% | 45 | 0·29% | 1·31 | (0·89, 1·94) | 0·1712 |
| **F43·1** | **Post-traumatic stress disorder (PTSD)** | 126 | 0·82% | 106 | 0·69% | 1·19 | (0·92, 1·54) | 0·1892 |
| **F43·2** | **Adjustment disorders** | 280 | 1·83% | 195 | 1·28% | 1·44 | (1·20, 1·74) | 0·0001* |
| **F60-F69** | **Disorders of adult personality and behavior** | 331 | 2·16% | 276 | 1·80% | 1·20 | (1·02, 1·41) | 0·0241 |
| **F60·3** | **Borderline personality disorder** | 59 | 0·39% | 53 | 0·35% | 1·11 | (0·77, 1·61) | 0·5821 |
| **F64** | **Gender identity disorders** | 13 | 0·09% | 11 | 0·07% | 1·18 | (0·53, 2·64) | 0·6965 |
| **F70-F79** | **Intellectual Disabilities** | 45 | 0·29% | 42 | 0·27% | 1·07 | (0·70, 1·63) | 0·7603 |
| **F84·0** | **Autistic disorder** | 184 | 1·20% | 153 | 1·00% | 1·21 | (0·97, 1·50) | 0·0897 |
| **F84·5** | **Asperger's syndrome** | 40 | 0·26% | 34 | 0·22% | 1·18 | (0·74, 1·86) | 0·4953 |
| **F90** | **Attention-deficit hyperactivity disorders** | 1181 | 7·72% | 894 | 5·85% | 1·35 | (1·23, 1·47) | <0·0008* |
| **F91** | **Conduct disorders** | 633 | 4·14% | 477 | 3·12% | 1·34 | (1·19, 1·51) | <0·0008* |
| **MEDICAL DIAGNOSES** | |  |  |  |  |  |  |  |
| **J45** | **Asthma** | 1172 | 7·66% | 878 | 5·74% | 1·36 | (1·24, 1·49) | <0·0008* |
| **E00-E07** | **Disorders of thyroid gland** | 70 | 0·46% | 62 | 0·41% | 1·13 | (0·80, 1·59) | 0·4954 |
| **E08-E13** | **Diabetes mellitus** | 88 | 0·58% | 58 | 0·38% | 1·52 | (1·09, 2·12) | 0·0134 |
| **G40** | **Epilepsy and recurrent seizures** | 130 | 0·85% | 105 | 0·69% | 1·24 | (0·96, 1·61) | 0·102 |
| **G47** | **Sleep disorders** | 562 | 3·67% | 419 | 2·74% | 1·35 | (1·19, 1·54) | <0·0008* |
| **G47·0** | **Insomnia** | 154 | 1·01% | 99 | 0·65% | 1·56 | (1·21, 2·01) | 0·0006* |
| **G43** | **Migraine** | 200 | 1·31% | 111 | 0·73% | 1·81 | (1·44, 2·29) | <0·0008* |

**Supplementary Table 3.** Prevalence and odds ratios of medical/psychiatric/socioeconomic characteristics in patients 13-21 with or without COVID-19 among SOSH+ patients**.** Statistical comparisons are represented by odds ratios (OR) with 95% confidence intervals (CI)**.** Significant p-values are indicated with an asterisk (*)**.**

| **ICD-10 Code** | **Characteristic** | **SI/SA(+), COVID-19(+)** | | **SI/SA(+), COVID-19(-)** | | **OR** | **95%CI** | **P value** |
| --- | --- | --- | --- | --- | --- | --- | --- | --- |
|  |  | **N** | **%** | **N** | **%** |  |  |  |
| **SOCIOECONOMIC DIAGNOSES** | |  |  |  |  |  |  |  |
| **Z55** | **Problems related to education and literacy** | 245 | 1·58% | 344 | 2·22% | 0·71 | (0·60, 0·83) | <0·0008* |
| **Z59** | **Problems related to housing and economic circumstances** | 114 | 0·74% | 99 | 0·64% | 1·15 | (0·88, 1·51) | 0·3071 |
| **Z60** | **Problems related to social environment** | 176 | 1·14% | 209 | 1·35% | 0·84 | (0·69, 1·03) | 0·0907 |
| **Z60.4** | **Social exclusion and rejection** | 73 | 0·47% | 59 | 0·38% | 1·24 | (0·88, 1·75) | 0·2249 |
| **Z62** | **Problems related to upbringing** | 960 | 6·20% | 1147 | 7·41% | 0·83 | (0·76, 0·90) | <0·0008* |
| **Z62·2** | **Upbringing away from parents** | 166 | 1·07% | 213 | 1·38% | 0·78 | (0·63, 0·95) | 0·0153 |
| **Z62.81** | **Personal history of abuse in childhood** | 585 | 3·78% | 639 | 4·13% | 0·91 | (0·81, 1·02) | 0·1153 |
| **Z62.810** | **Personal history of physical and sexual abuse in childhood** | 416 | 2·69% | 478 | 3·09% | 0·87 | (0·76, 0·99) | 0·0352 |
| **Z62.811** | **Personal history of psychological abuse in childhood** | 97 | 0·63% | 220 | 1·42% | 0·44 | (0·34, 0·56) | <0·0008* |
| **Z62.812** | **Personal history of neglect in childhood** | 45 | 0·29% | 61 | 0·39% | 0·74 | (0·50, 1·08) | 0·121 |
| **Z62.819** | **Personal history of unspecified abuse in childhood** | 128 | 0·83% | 50 | 0·32% | 2·57 | (1·85, 3·57) | <0·0008* |
| **Z62·82** | **Parent-child conflict** | 293 | 1·89% | 447 | 2·89% | 0·65 | (0·56, 0·75) | <0·0008* |
| **Z63** | **Other problems related to primary support group, including family circumstances** | 788 | 5·09% | 687 | 4·44% | 1·15 | (1·04, 1·28) | 0·0071 |
| **Z65** | **Problems related to other psychosocial circumstances** | 216 | 1·40% | 275 | 1·78% | 0·78 | (0·65, 0·94) | 0·0074 |
| **Z81** | **Family history of mental and behavioral disorders** | 757 | 4·89% | 958 | 6·19% | 0·78 | (0·71, 0·86) | <0·0008* |
| **Z91.5** | **Personal history of self-harm** | 1478 | 9·55% | 1674 | 10·82% | 0·87 | (0·81, 0·94) | 0·0003* |
| **Z91.52** | **Personal history of nonsuicidal self-harm** | 168 | 1·09% | 295 | 1·91% | 0·56 | (0·47, 0·68) | <0·0008* |
| **PSYCHIATRIC DIAGNOSES** | |  |  |  |  |  |  |  |
| **F10-F19** | **Mental and behavioral disorders due to psychoactive substance use** | 1041 | 6·73% | 1208 | 7·81% | 0·85 | (0·78, 0·93) | 0·0003* |
| **F10** | **Alcohol related disorders** | 159 | 1·03% | 244 | 1·58% | 0·65 | (0·53, 0·79) | <0·0008* |
| **F12** | **Cannabis related disorders** | 596 | 3·85% | 708 | 4·58% | 0·84 | (0·75, 0·93) | 0·0016 |
| **F17** | **Nicotine dependence** | 412 | 2·66% | 403 | 2·60% | 1·02 | (0·89, 1·18) | 0·7622 |
| **F20-F29** | **Schizophrenia, schizotypal, delusional, and other non-mood psychotic disorders** | 361 | 2·33% | 319 | 2·06% | 1·13 | (0·97, 1·32) | 0·1034 |
| **F20** | **Schizophrenia** | 96 | 0·62% | 75 | 0·48% | 1·28 | (0·95, 1·74) | 0·1081 |
| **F22** | **Delusional disorders** | 49 | 0·32% | 44 | 0·28% | 1·11 | (0·74, 1·67) | 0·6162 |
| **F25** | **Schizoaffective disorders** | 26 | 0·17% | 31 | 0·20% | 0·84 | (0·50, 1·41) | 0·5184 |
| **F29** | **Unspecified psychosis not due to a substance or known physiological condition** | 205 | 1·32% | 185 | 1·20% | 1·11 | (0·91, 1·36) | 0·3128 |
| **F30-F39** | **Mood [affective] disorders** | 5281 | 34·13% | 5929 | 38·32% | 0·83 | (0·80, 0·87) | <0·0008* |
| **F31** | **Bipolar disorder** | 497 | 3·21% | 361 | 2·33% | 1·39 | (1·21, 1·59) | <0·0008* |
| **F32** | **Depressive episode** | 4145 | 26·79% | 4616 | 29·83% | 0·86 | (0·82, 0·90) | <0·0008* |
| **F33** | **Major depressive disorder, recurrent** | 1170 | 7·56% | 1627 | 10·52% | 0·70 | (0·64, 0·75) | <0·0008* |
| **F34.1** | **Dysthymic disorder** | 100 | 0·65% | 126 | 0·81% | 0·79 | (0·61, 1·03) | 0·0829 |
| **F40-F48** | **Anxiety, dissociative, stress-related, somatoform and other nonpsychotic mental disorders** | 4817 | 31·13% | 4935 | 31·89% | 0·97 | (0·92, 1·01) | 0·1492 |
| **F41·0** | **Panic disorder [episodic paroxysmal anxiety]** | 348 | 2·25% | 353 | 2·28% | 0·99 | (0·85, 1·14) | 0·8589 |
| **F41·1** | **Generalized anxiety disorder** | 1059 | 6·84% | 1220 | 7·88% | 0·86 | (0·79, 0·93) | 0·0005* |
| **F41·9** | **Anxiety disorder, unspecified** | 2967 | 19·18% | 2656 | 17·17% | 1·14 | (1·08, 1·21) | <0·0008* |
| **F42** | **Obsessive-compulsive disorder** | 214 | 1·38% | 257 | 1·66% | 0·83 | (0·69, 1·00) | 0·0458 |
| **F43·1** | **Post-traumatic stress disorder (PTSD)** | 896 | 5·79% | 915 | 5·91% | 0·98 | (0·89, 1·08) | 0·6584 |
| **F43·2** | **Adjustment disorders** | 1038 | 6·71% | 981 | 6·34% | 1·06 | (0·97, 1·16) | 0·1908 |
| **F60-F69** | **Disorders of adult personality and behavior** | 836 | 5·40% | 839 | 5·42% | 1·00 | (0·90, 1·10) | 0·9452 |
| **F60·3** | **Borderline personality disorder** | 194 | 1·25% | 176 | 1·14% | 1·10 | (0·90, 1·36) | 0·3523 |
| **F64** | **Gender identity disorders** | 173 | 1·12% | 243 | 1·57% | 0·71 | (0·58, 0·86) | 0·0006* |
| **F70-F79** | **Intellectual Disabilities** | 140 | 0·90% | 108 | 0·70% | 1·30 | (1·01, 1·67) | 0·0416 |
| **F84·0** | **Autistic disorder** | 450 | 2·91% | 446 | 2·88% | 1·01 | (0·88, 1·15) | 0·9004 |
| **F84·5** | **Asperger's syndrome** | 72 | 0·47% | 60 | 0·39% | 1·20 | (0·85, 1·69) | 0·3 |
| **F90** | **Attention-deficit hyperactivity disorders** | 2584 | 16·70% | 2138 | 13·82% | 1·25 | (1·18, 1·33) | <0·0008* |
| **F91** | **Conduct disorders** | 1310 | 8·47% | 1039 | 6·71% | 1·28 | (1·18, 1·40) | <0·0008* |
| **MEDICAL DIAGNOSES** | |  |  |  |  |  |  |  |
| **J45** | **Asthma** | 1870 | 12·09% | 1570 | 10·15% | 1·22 | (1·13, 1·31) | <0·0008* |
| **E00-E07** | **Disorders of thyroid gland** | 201 | 1·30% | 177 | 1·14% | 1·14 | (0·93, 1·39) | 0·2163 |
| **E08-E13** | **Diabetes mellitus** | 232 | 1·50% | 141 | 0·91% | 1·66 | (1·34, 2·04) | <0·0008* |
| **G40** | **Epilepsy and recurrent seizures** | 243 | 1·57% | 193 | 1·25% | 1·26 | (1·04, 1·53) | 0·016 |
| **G47** | **Sleep disorders** | 1300 | 8·40% | 1140 | 7·37% | 1·15 | (1·06, 1·25) | 0·0008* |
| **G47·0** | **Insomnia** | 568 | 3·67% | 557 | 3·60% | 1·02 | (0·91, 1·15) | 0·7513 |
| **G43** | **Migraine** | 486 | 3·14% | 425 | 2·75% | 1·15 | (1·01, 1·31) | 0·04 |

**Supplementary Table 4.** Prevalence and odds ratios of medical/psychiatric/socioeconomic characteristics in patients 13-21 with or without COVID-19 among SI/SA+ patients**.** Statistical comparisons are represented by odds ratios (OR) with 95% confidence intervals (CI)**.** Significant p-values are indicated with an asterisk (*)**.**

| **ICD-10 Code** | **Characteristic** | **COVID-19(+), SOSH(+)** | | **COVID-19(-), SOSH(-)** | | **OR** | **95%CI** | **P value** |
| --- | --- | --- | --- | --- | --- | --- | --- | --- |
|  |  | **N** | **%** | **N** | **%** |  |  |  |
| **PSYCHIATRIC DIAGNOSES** | |  |  |  |  |  |  |  |
| **F40-F48** | **Anxiety, dissociative, stress-related, somatoform and other nonpsychotic mental disorders** | 281 | 20·69% | 46 | 3·39% | 7·44 | (5·39, 10·27) | *<0·0008 |
| **F41·9** | **Anxiety disorder, unspecified** | 140 | 10·31% | 17 | 1·25% | 9·07 | (5·45, 15·09) | *<0·0008 |
| **F84·0** | **Autistic disorder** | 47 | 3·46% | 19 | 1·40% | 2·53 | (1·47, 4·33) | 0·0008 |
| **F90** | **Attention-deficit hyperactivity disorders** | 293 | 21·58% | 47 | 3·46% | 7·67 | (5·58, 10·55) | *<0·0008 |
| **F91** | **Conduct disorders** | 164 | 12·08% | 21 | 1·55% | 8·74 | (5·52, 13·87) | *<0·0008 |
| **MEDICAL DIAGNOSES** | |  |  |  |  |  |  |  |
| **J45** | **Asthma** | 244 | 17·97% | 113 | 8·32% | 2·41 | (1·90, 3·06) | *<0·0008 |
| **G40** | **Epilepsy and recurrent seizures** | 29 | 2·14% | 22 | 1·62% | 1·33 | (0·76, 2·32) | 0·3289 |
| **G47** | **Sleep disorders** | 145 | 10·68% | 57 | 4·20% | 2·73 | (1·99, 3·74) | *<0·0008 |
| **G43** | **Migraine** | 13 | 0·96% | 14 | 1·03% | 0·93 | (0·43, 1·98) | 0·8571 |

**Supplementary Table 5·** Prevalence and odds ratios of medical/psychiatric/socioeconomic characteristics in patients 0-12 with COVID-19 and SOSH compared to patients without COVID-19 and without SOSH· Statistical comparisons are represented by odds ratios (OR) with 95% confidence intervals (CI)· Significant p-values are indicated with an asterisk (*)·

| **ICD-10 Code** | **Characteristic** | **COVID-19(+), SI/SA(+)** | | **COVID-19(-), SI/SA(-)** | | **OR** | **95%CI** | **P value** |
| --- | --- | --- | --- | --- | --- | --- | --- | --- |
|  |  | **N** | **%** | **N** | **%** |  |  |  |
| **PSYCHIATRIC DIAGNOSES** | |  |  |  |  |  |  |  |
| **F40-F48** | **Anxiety, dissociative, stress-related, somatoform and other nonpsychotic mental disorders** | 273 | 20·94% | 36 | 2·76% | 9·33 | (6·53, 13·33) | *<0·0008 |
| **F41·9** | **Anxiety disorder, unspecified** | 138 | 10·58% | 15 | 1·15% | 10·17 | (5·93, 17·43) | *<0·0008 |
| **F43·2** | **Adjustment disorders** | 80 | 6·13% | 11 | 0·84% | 7·68 | (4·07, 14·50) | *<0·0008 |
| **F84·0** | **Autistic disorder** | 42 | 3·22% | 16 | 1·23% | 2·68 | (1·50, 4·79) | 0·0009 |
| **F90** | **Attention-deficit hyperactivity disorders** | 293 | 22·47% | 34 | 2·61% | 10·83 | (7·52, 15·59) | *<0·0008 |
| **F91** | **Conduct disorders** | 163 | 12·50% | 17 | 1·30% | 10·82 | (6·52, 17·94) | *<0·0008 |
| **MEDICAL DIAGNOSES** | |  |  |  |  |  |  |  |
| **J45** | **Asthma** | 229 | 17·56% | 115 | 8·82% | 2·20 | (1·73, 2·80) | *<0·0008 |
| **G40** | **Epilepsy and recurrent seizures** | 24 | 1·84% | 17 | 1·30% | 1·42 | (0·76, 2·65) | 0·2762 |
| **G47** | **Sleep disorders** | 140 | 10·74% | 51 | 3·91% | 2·95 | (2·12, 4·11) | *<0·0008 |

Supplementary Table 6**.** Prevalence and odds ratios of medical/psychiatric/socioeconomic characteristics in patients 0-12 with COVID-19 and SI/SA compared to patients without COVID-19 and without SI/SA**.** Statistical comparisons are represented by odds ratios (OR) with 95% confidence intervals (CI)**.** Significant p-values are indicated with an asterisk (*)**.**

| **ICD-10 Code** | **Characteristic** | **COVID-19(+), SI/SA(+)** | | **COVID-19(-), SI/SA(-)** | | **OR** | **95%CI** | **P value** |
| --- | --- | --- | --- | --- | --- | --- | --- | --- |
|  |  | **N** | **%** | **N** | **%** |  |  |  |
| **SOCIOECONOMIC DIAGNOSES** | |  |  |  |  |  |  |  |
| **Z55** | **Problems related to education and literacy** | 80 | 0·52% | 41 | 0·27% | 1·96 | (1·34, 2·85) | 0·0005* |
| **Z60** | **Problems related to social environment** | 25 | 0·16% | 14 | 0·09% | 1·79 | (0·93, 3·44) | 0·0818 |
| **Z62** | **Problems related to upbringing** | 149 | 0·97% | 43 | 0·28% | 3·49 | (2·48, 4·90) | <0·0008* |
| **Z62·2** | **Upbringing away from parents** | 42 | 0·27% | 16 | 0·10% | 2·63 | (1·48, 4·68) | 0·0011 |
| **Z62·82** | **Parent-child conflict** | 54 | 0·35% | 13 | 0·08% | 4·17 | (2·27, 7·63) | <0·0008* |
| **Z63** | **Other problems related to primary support group, including family circumstances** | 146 | 0·95% | 50 | 0·33% | 2·94 | (2·13, 4·05) | <0·0008* |
| **Z65** | **Problems related to other psychosocial circumstances** | 62 | 0·41% | 18 | 0·12% | 3·45 | (2·04, 5·84) | <0·0008* |
| **Z81** | **Family history of mental and behavioral disorders** | 105 | 0·69% | 22 | 0·14% | 4·80 | (3·03, 7·60) | <0·0008* |
| **PSYCHIATRIC DIAGNOSES** | |  |  |  |  |  |  |  |
| **F30-F39** | **Mood [affective] disorders** | 668 | 4·37% | 135 | 0·88% | 5·13 | (4·26, 6·18) | <0·0008* |
| **F31** | **Bipolar disorder** | 75 | 0·49% | 14 | 0·09% | 5·38 | (3·04, 9·52) | <0·0008* |
| **F32** | **Depressive episode** | 460 | 3·01% | 95 | 0·62% | 4·96 | (3·97, 6·19) | <0·0008* |
| **F33** | **Major depressive disorder, recurrent** | 91 | 0·59% | 12 | 0·08% | 7·62 | (4·17, 13·92) | <0·0008* |
| **F40-F48** | **Anxiety, dissociative, stress-related, somatoform and other nonpsychotic mental disorders** | 1066 | 6·97% | 362 | 2·37% | 3·09 | (2·74, 3·49) | <0·0008* |
| **F41·0** | **Panic disorder [episodic paroxysmal anxiety]** | 42 | 0·27% | 12 | 0·08% | 3·51 | (1·85, 6·66) | 0·0001* |
| **F41·1** | **Generalized anxiety disorder** | 216 | 1·41% | 81 | 0·53% | 2·69 | (2·08, 3·48) | <0·0008* |
| **F41·9** | **Anxiety disorder, unspecified** | 577 | 3·77% | 188 | 1·23% | 3·15 | (2·67, 3·72) | <0·0008* |
| **F42** | **Obsessive-compulsive disorder** | 59 | 0·39% | 28 | 0·18% | 2·11 | (1·35, 3·31) | 0·0012 |
| **F43·1** | **Post-traumatic stress disorder (PTSD)** | 126 | 0·82% | 17 | 0·11% | 7·47 | (4·50, 12·39) | <0·0008* |
| **F43·2** | **Adjustment disorders** | 280 | 1·83% | 103 | 0·67% | 2·75 | (2·19, 3·45) | <0·0008* |
| **F60-F69** | **Disorders of adult personality and behavior** | 331 | 2·16% | 112 | 0·73% | 3·00 | (2·42, 3·72) | <0·0008* |
| **F60·3** | **Borderline personality disorder** | 59 | 0·39% | 13 | 0·08% | 4·55 | (2·50, 8·30) | <0·0008* |
| **F70-F79** | **Intellectual Disabilities** | 45 | 0·29% | 45 | 0·29% | 1·00 | (0·66, 1·51) | 1 |
| **F84·0** | **Autistic disorder** | 184 | 1·20% | 94 | 0·61% | 1·97 | (1·53, 2·53) | <0·0008* |
| **F84·5** | **Asperger's syndrome** | 40 | 0·26% | 17 | 0·11% | 2·36 | (1·34, 4·16) | 0·0031 |
| **F90** | **Attention-deficit hyperactivity disorders** | 1181 | 7·72% | 455 | 2·97% | 2·73 | (2·44, 3·05) | <0·0008* |
| **F91** | **Conduct disorders** | 633 | 4·14% | 187 | 1·22% | 3·49 | (2·96, 4·11) | <0·0008* |
| **MEDICAL DIAGNOSES** | |  |  |  |  |  |  |  |
| **J45** | **Asthma** | 1172 | 7·66% | 801 | 5·24% | 1·50 | (1·37, 1·65) | <0·0008* |
| **E00-E07** | **Disorders of thyroid gland** | 70 | 0·46% | 91 | 0·59% | 0·77 | (0·56, 1·05) | 0·0977 |
| **E08-E13** | **Diabetes mellitus** | 88 | 0·58% | 75 | 0·49% | 1·17 | (0·86, 1·60) | 0·3122 |
| **G40** | **Epilepsy and recurrent seizures** | 130 | 0·85% | 148 | 0·97% | 0·88 | (0·69, 1·11) | 0·2821 |
| **G47** | **Sleep disorders** | 562 | 3·67% | 309 | 2·02% | 1·85 | (1·61, 2·13) | <0·0008* |
| **G47·0** | **Insomnia** | 154 | 1·01% | 49 | 0·32% | 3·16 | (2·29, 4·37) | <0·0008* |
| **G43** | **Migraine** | 200 | 1·31% | 117 | 0·76% | 1·72 | (1·37, 2·16) | <0·0008* |

Supplementary Table 7**.** Prevalence and odds ratios of medical/psychiatric/socioeconomic characteristics in patients 13-21 with COVID-19 and SOSH compared to patients without COVID-19 and without SOSH**.** Statistical comparisons are represented by odds ratios (OR) with 95% confidence intervals (CI)**.** Significant p-values are indicated with an asterisk (*)**.**

| **ICD-10 Code** | **Characteristic** |  | |  | | **OR** | **95%CI** | **P value** |
| --- | --- | --- | --- | --- | --- | --- | --- | --- |
|  |  | **N** | **%** | **N** | **%** |  |  |  |
| **SOCIOECONOMIC DIAGNOSES** | |  |  |  |  |  |  |  |
| **Z55** | **Problems related to education and literacy** | 245 | 1·58% | 94 | 0·61% | 2·63 | (2·07, 3·34) | <0·0008* |
| **Z59** | **Problems related to housing and economic circumstances** | 114 | 0·74% | 36 | 0·23% | 3·18 | (2·19, 4·63) | <0·0008* |
| **Z60** | **Problems related to social environment** | 176 | 1·14% | 19 | 0·12% | 9·36 | (5·83, 15·03) | <0·0008* |
| **Z62** | **Problems related to upbringing** | 960 | 6·20% | 114 | 0·74% | 8·91 | (7·33, 10·84) | <0·0008* |
| **Z62·2** | **Upbringing away from parents** | 166 | 1·07% | 50 | 0·32% | 3·35 | (2·44, 4·59) | <0·0008* |
| **Z62.81** | **Personal history of abuse in childhood** | 585 | 3·78% | 36 | 0·23% | 16·85 | (12·02, 23·61) | <0·0008* |
| **Z62.810** | **Personal history of physical and sexual abuse in childhood** | 416 | 2·69% | 23 | 0·15% | 18·56 | (12·19, 28·26) | <0·0008* |
| **Z62·82** | **Parent-child conflict** | 293 | 1·89% | 34 | 0·22% | 8·76 | (6·14, 12·51) | <0·0008* |
| **Z63** | **Other problems related to primary support group, including family circumstances** | 788 | 5·09% | 92 | 0·59% | 8·97 | (7·22, 11·15) | <0·0008* |
| **Z65** | **Problems related to other psychosocial circumstances** | 216 | 1·40% | 40 | 0·26% | 5·46 | (3·90, 7·66) | <0·0008* |
| **Z81** | **Family history of mental and behavioral disorders** | 757 | 4·89% | 83 | 0·54% | 9·54 | (7·60, 11·98) | <0·0008* |
| **Z91.5** | **Personal history of self-harm** | 1478 | 9·55% | 51 | 0·33% | 31·94 | (24·13, 42·26) | <0·0008* |
| **PSYCHIATRIC DIAGNOSES** | |  |  |  |  |  |  |  |
| **F10-F19** | **Mental and behavioral disorders due to psychoactive substance use** | 1041 | 6·73% | 129 | 0·83% | 8·58 | (7·14, 10·32) | <0·0008* |
| **F10** | **Alcohol related disorders** | 159 | 1·03% | 18 | 0·12% | 8·91 | (5·47, 14·52) | <0·0008* |
| **F12** | **Cannabis related disorders** | 596 | 3·85% | 68 | 0·44% | 9·08 | (7·05, 11·68) | <0·0008* |
| **F17** | **Nicotine dependence** | 412 | 2·66% | 41 | 0·26% | 10·30 | (7·46, 14·20) | <0·0008* |
| **F20-F29** | **Schizophrenia, schizotypal, delusional, and other non-mood psychotic disorders** | 361 | 2·33% | 24 | 0·16% | 15·38 | (10·17, 23·26) | <0·0008* |
| **F29** | **Unspecified psychosis not due to a substance or known physiological condition** | 205 | 1·32% | 12 | 0·08% | 17·30 | (9·66, 30·98) | <0·0008* |
| **F30-F39** | **Mood [affective] disorders** | 5281 | 34·13% | 636 | 4·11% | 12·09 | (11·09, 13·17) | <0·0008* |
| **F31** | **Bipolar disorder** | 497 | 3·21% | 37 | 0·24% | 13·84 | (9·91, 19·35) | <0·0008* |
| **F32** | **Depressive episode** | 4145 | 26·79% | 475 | 3·07% | 11·55 | (10·47, 12·74) | <0·0008* |
| **F33** | **Major depressive disorder, recurrent** | 1170 | 7·56% | 116 | 0·75% | 10·83 | (8·94, 13·12) | <0·0008* |
| **F34.1** | **Dysthymic disorder** | 100 | 0·65% | 28 | 0·18% | 3·59 | (2·36, 5·46) | <0·0008* |
| **F40-F48** | **Anxiety, dissociative, stress-related, somatoform and other nonpsychotic mental disorders** | 4817 | 31·13% | 1206 | 7·79% | 5·35 | (5·00, 5·72) | <0·0008* |
| **F41·0** | **Panic disorder [episodic paroxysmal anxiety]** | 348 | 2·25% | 57 | 0·37% | 6·22 | (4·70, 8·24) | <0·0008* |
| **F41·1** | **Generalized anxiety disorder** | 1059 | 6·84% | 247 | 1·60% | 4·53 | (3·94, 5·21) | <0·0008* |
| **F41·9** | **Anxiety disorder, unspecified** | 2967 | 19·17% | 720 | 4·65% | 4·86 | (4·47, 5·29) | <0·0008* |
| **F42** | **Obsessive-compulsive disorder** | 214 | 1·38% | 45 | 0·29% | 4·81 | (3·48, 6·64) | <0·0008* |
| **F43·1** | **Post-traumatic stress disorder (PTSD)** | 896 | 5·79% | 91 | 0·59% | 10·39 | (8·36, 12·91) | <0·0008* |
| **F43·2** | **Adjustment disorders** | 1038 | 6·71% | 221 | 1·43% | 4·96 | (4·28, 5·75) | <0·0008* |
| **F60-F69** | **Disorders of adult personality and behavior** | 836 | 5·40% | 188 | 1·21% | 4·64 | (3·96, 5·45) | <0·0008* |
| **F60·3** | **Borderline personality disorder** | 194 | 1·25% | 15 | 0·10% | 13·08 | (7·73, 22·14) | <0·0008* |
| **F64** | **Gender identity disorders** | 173 | 1·12% | 52 | 0·34% | 3·35 | (2·46, 4·58) | <0·0008* |
| **F70-F79** | **Intellectual Disabilities** | 140 | 0·90% | 76 | 0·49% | 1·85 | (1·40, 2·45) | <0·0008* |
| **F84·0** | **Autistic disorder** | 450 | 2·91% | 182 | 1·18% | 2·52 | (2·12, 2·99) | <0·0008* |
| **F84·5** | **Asperger's syndrome** | 72 | 0·47% | 19 | 0·12% | 3·80 | (2·29, 6·31) | <0·0008* |
| **F90** | **Attention-deficit hyperactivity disorders** | 2584 | 16·70% | 760 | 4·91% | 3·88 | (3·57, 4·22) | <0·0008* |
| **F91** | **Conduct disorders** | 1310 | 8·47% | 230 | 1·49% | 6·13 | (5·32, 7·07) | <0·0008* |
| **MEDICAL DIAGNOSES** | |  |  |  |  |  |  |  |
| **J45** | **Asthma** | 1870 | 12·08% | 1246 | 8·05% | 1·57 | (1·46, 1·69) | <0·0008* |
| **E00-E07** | **Disorders of thyroid gland** | 201 | 1·30% | 199 | 1·29% | 1·01 | (0·83, 1·23) | 0·9265 |
| **E08-E13** | **Diabetes mellitus** | 232 | 1·50% | 153 | 0·99% | 1·52 | (1·24, 1·87) | <0·0008* |
| **G40** | **Epilepsy and recurrent seizures** | 243 | 1·57% | 225 | 1·45% | 1·08 | (0·90, 1·30) | 0·4093 |
| **G47** | **Sleep disorders** | 1300 | 8·40% | 592 | 3·83% | 2·31 | (2·09, 2·55) | <0·0008* |
| **G47·0** | **Insomnia** | 568 | 3·67% | 117 | 0·76% | 5·00 | (4·09, 6·11) | <0·0008* |
| **G43** | **Migraine** | 486 | 3·14% | 349 | 2·26% | 1·41 | (1·22, 1·62) | <0·0008* |

Supplementary Table 8**.** Prevalence and odds ratios of medical/psychiatric/socioeconomic characteristics in patients 13-21 with COVID-19 and SI/SA compared to patients without COVID-19 and without SI/SA**.** Statistical comparisons are represented by odds ratios (OR) with 95% confidence intervals (CI)**.** Significant p-values are indicated with an asterisk (*)**.**

**Appendix**

Altman DG and Bland M**.** How to obtain the P value from a confidence interval**.** *BMJ* 2011; **343**: d2304. doi: [https://doi·org/10·1136/bmj·d2304](https://doi.org/10.1136/bmj.d2304)

Higgins J and Thomas J**.** Cochrane Handbook for Systematic Reviews of Interventions**.** [https://training·cochrane·org/handbook/current](https://training.cochrane.org/handbook/current)· 2 April 2024**.**
